# The selective pressure induced by malaria vaccines on *Plasmodium falciparum*

**DOI:** 10.1101/2024.05.22.24307679

**Authors:** Thiery Masserey, Tamsin Lee, Aurélien Cavelan, Daniel E. Neafsey, Josephine Malinga, Melissa A Penny

**Affiliations:** Swiss Tropical and Public Health Institute, Allschwil, Switzerland; University of Basel, Basel, Switzerland; Harvard T. H. Chan School of Public Health, Boston, USA; Broad Institute of MIT and Harvard, Cambridge, USA; University of Western Australia, Perth, Australia; Telethon Kids Institute, Nedlands, Australia

## Abstract

Antigens of *Plasmodium falciparum* targeted by most current and future vaccines are generally not conserved. There are limited studies estimating the risk of parasite selection with vaccines. We adapted an individual-based model of malaria to assess which conditions favour genotypes having some degree of vaccine resistance and estimate the impact of resistance spread on vaccine effectiveness. Even parasite genotypes with low degrees of vaccine resistance are likely to spread relatively quickly if vaccines are implemented in children and even faster in a broader population. However, only highly resistant genotypes could strongly reduce vaccine effectiveness. These results highlight that it is essential to understand the degree to which genotypes exhibit reduced sensitivity to vaccines and monitor genotype frequency and vaccine effectiveness along with vaccine deployment. If some genotypes exhibit reduced efficacy to vaccines, our results further suggest that polyvalent or combination vaccines should be considered to limit resistance spread.

## Main

Malaria caused by *Plasmodium falciparum* continues to cause a substantial health burden globally^4^. Developing new technologies, such as vaccines, will be crucial in reducing this burden^4^. In 2022, the World Health Organization (WHO) outlined two use cases for malaria vaccines in their documents that guide vaccine development^5^. The first use case aims to deploy vaccines that reduce morbidity and mortality in vulnerable populations, primarily children under five^5,6^. These vaccines could be an anti-infective vaccine (AIV), preventing infection by targeting the pre-erythrocyte stage of the parasite, or a blood-stage vaccine (BSV), limiting the parasite blood-stage multiplication^5,6^ (Fig. 1A). The second use case aims to deploy vaccines to the entire population or solely to adults to diminish malaria transmission^5,6^. These vaccines are likely to consist of a highly efficient AIV or an AIV combined with a transmission-blocking vaccine (TBV) targeting the sexual stage of the parasites^5,6^ (Fig. 1A).

**Figure 1:**
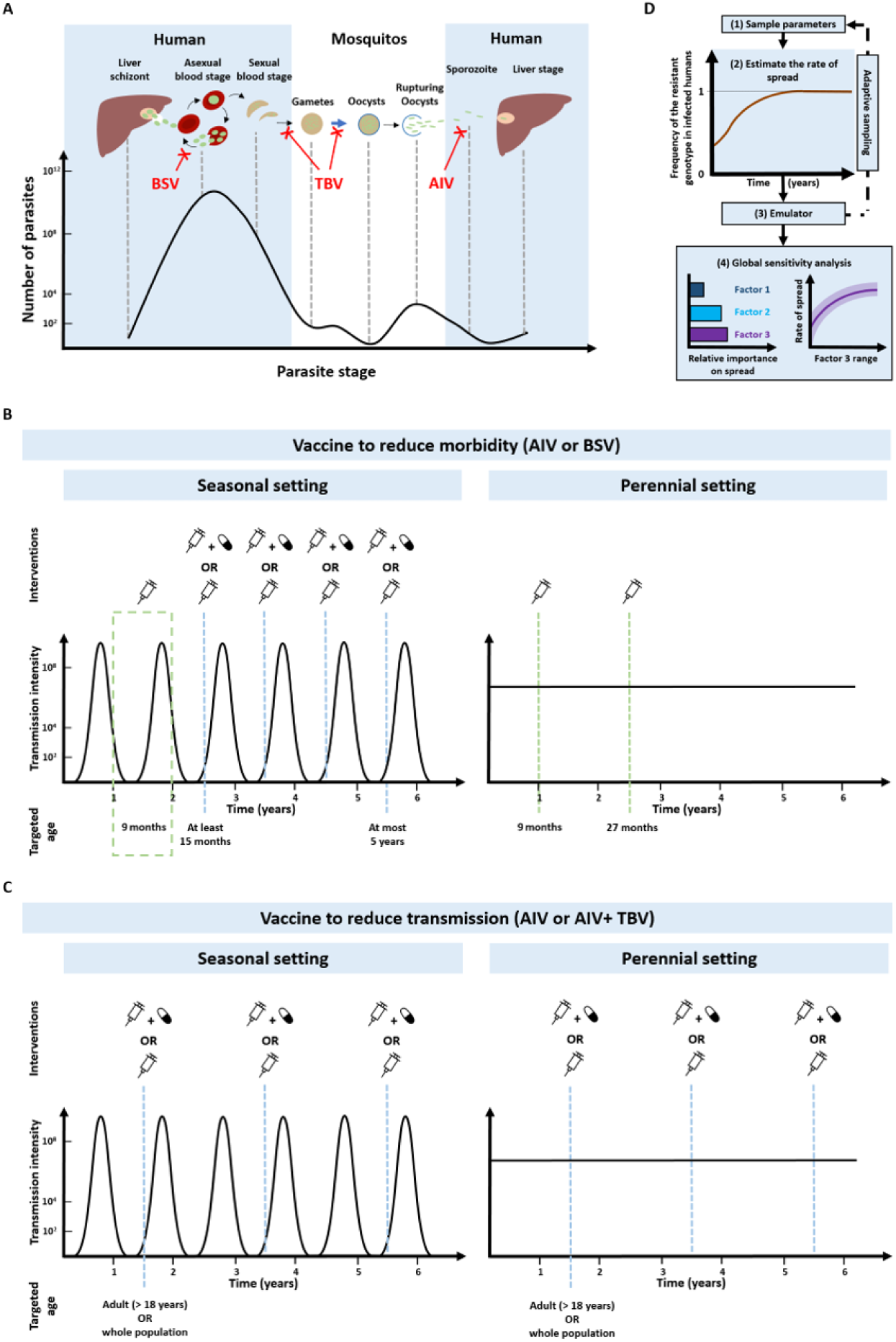
Vaccine actions, deployment and analysis workflow. (**A**) *P. falciparum* density across its life cycle^44^, and the impact of AIVs, BSVs, and TBVs. The blue arrow indicates when recombination occurs between the gametocyte genomes. (**B**) Strategies used to deploy an AIV or BSV to reduce morbidity. In the seasonal transmission setting, the primary vaccination series was delivered to children at nine months of age. Then, vaccinated children older than 15 months received a booster dose with or without a drug two months before the transmission peak for four years (methods). In the perennial transmission setting, the primary vaccination series was delivered to children at nine months of age with a booster dose 18 months later. (**C**) Strategies used to deploy an AIV or AIV+TBV to reduce transmission. The AIV was deployed with or without a drug to the whole population or only adults. The vaccine was deployed through mass administration with a booster dose every two years (methods). The deployment occurred two months before the transmission peak in the seasonal setting. (**D**) We systematically assessed the relative impact of factors listed in Supplementary Table 8 on the spread of vaccine-resistant parasites through global sensitivity analysis (methods). This involved (1) randomly sampling combinations of parameters, (2) assessing the rate of spread (as the selection coeffcient, methods) of resistant genotypes for each parameter combination, (3) training an emulator to learn the relationship between the input and the rate of spread with iterative fitting improvements through adaptive sampling, (4) performing global sensitivity using the emulator. Global sensitivity analysis estimates the relative influence of each factor on the rate of spread and the 25th, 50th, and 75th quantiles of the estimated selection coefficient across each parameter range.

In 2021, the WHO recommended the broad paediatric use of the first AIV, RTS,S/AS01, conferring partial protection against clinical malaria^7,8^. More recently a second AIV, R21/Matrix-M, was recommended by WHO^9^. Moreover, many BSVs (such as RH5 and AMA1-RON2) and TBVs (like Pfs25 and Pfs230) are reaching advanced steps of development. However, RTS,S/AS01 and most vaccines in development are based on a single parasite genotype^10^. Several studies have reported that real-world parasites have different alleles of the antigen targeted by most vaccines^1–3^. Furthermore, studies have reported that some of these vaccines, including RTS,S/AS01, may have reduced efficacy against parasites with an antigen allele other than the one included in the vaccine^11–18^. Previously, for other pathogens, differences in vaccine efficacy between genotypes resulted in the spread of vaccine-resistant genotypes^19–27^.

Only a few studies have examined the potential risk of vaccine resistance for *P. falciparum*. We still do not know the extent of antigen diversity for RTS,S/AS01 and most vaccines under development, and how much vaccine efficacy varies between genotypes. Furthermore, we do not know under which conditions such a genotype would spread and how it would impact vaccine effectiveness. Mathematical models have previously assessed the spread of vaccine-resistant genotypes for other pathogens^28–43^. However, no model has focused on *P. falciparum* and its complex life cycle, nor evaluated how the spread of vaccine-resistant genotypes can be influenced by different vaccine actions (prevent infection, morbidity or transmission), vaccine proprieties (efficacy and duration of protection), and deployment strategies,, and how this spread could impact vaccine efficacy at the population level.

Given the history of vaccine resistance for other pathogens, the increased investment in malaria vaccine development, and the ongoing implementation of RTS,S/AS01 and soon R21, assessing the potential risk of vaccine resistance for *P. falciparum* is essential. In this study, we adapted an existing individual-based model of malaria, which includes a mechanistic within-host model, explicitly model mechanisms of immunity acquisition and vaccine effects and track multiple genotypes (see methods). Using this model, we assessed the rate of spread of a genotype resistant to an AIV or a BSV administered to children to prevent morbidity and mortality (Fig. 1B) and parasites resistant to an AIV delivered to the whole population or only adults to reduce malaria transmission (Fig. 1C). We then assessed which conditions favour the rate of spread of vaccine-resistant genotypes, including vaccine actions, proprieties, and deployment strategies, and we estimated how the spread of such resistant genotypes could impact the effectiveness of vaccines.

## Results

### Influence of factors on resistance spread

Through global sensitivity analyses, we systematically assessed which biological, epidemiological, vaccine, health system, and deployment factors (Supplementary Table 8) favour the spread of vaccine-resistant parasites for each malaria vaccine type and use case (Fig. 1D, and methods). For all scenarios, the degree of resistance of the parasites to the vaccine (defined as the relative decrease in the initial vaccine efficacy of the resistant genotype compared with the sensitive one) has the greatest influence on the rate of spread (estimated through the selection coefficient, see methods) (Fig. 2A). The estimated rate of spread of resistant parasites accelerates strongly with higher degrees of vaccine resistance (Fig. 2B), highlighting the importance of evaluating the degree of resistance exhibited by genotypes in real world settings.

**Figure 2:**
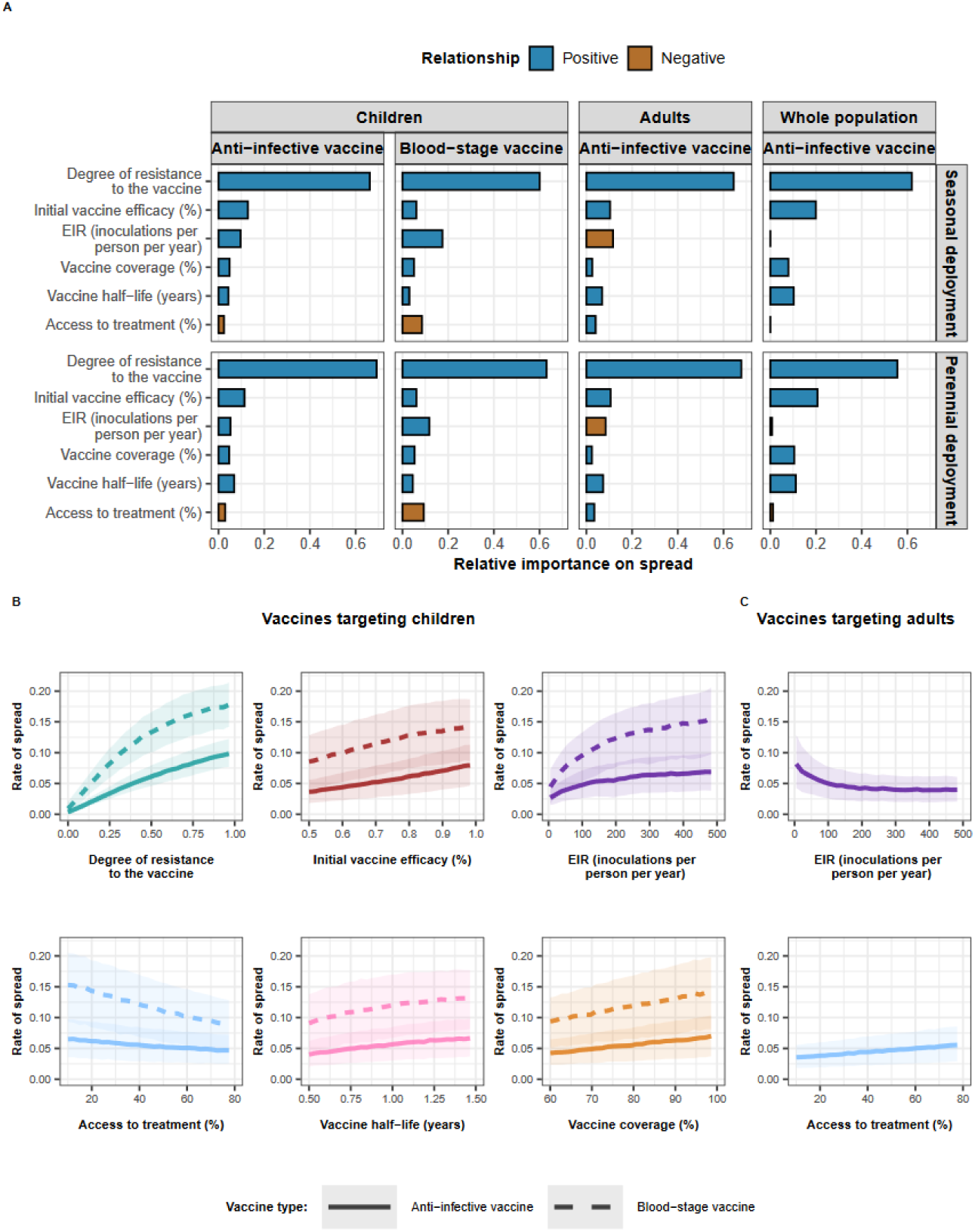
The drivers of the spread of vaccine-resistant genotypes for each deployment strategy and vaccine type. (**A**) The relative contribution of each factor was assessed through global sensitivity analysis when an AIV or a BSV was deployed to children in a perennial or seasonal setting, and when an AIV was deployed to the whole population or only adults. Factors in blue and brown indicate a positive and negative relationship with the spread, respectively. The parameter ranges and definitions are described in Supplementary Table 8. (**B**) Influence of each factor on the rate of spread (estimated trough the selection coefficient) of a vaccine-resistant genotype when an AIV or BSV was implemented in children in a seasonal setting. Curves and shaded areas represent the median and interquartile range of the rate of spread of a genotype resistant to an AIV (solid curve) or a BSV (dashed curve) estimated during the global sensitivity analysis over the parameter range described in Table 8. (**C**) Influence of EIR and access to treatment on the spread of a genotype resistant to an AIV deployed to adults in a seasonal setting. Curves and shaded areas represent the median and interquartile range of the rate of spread of a resistant genotype estimated during the global sensitivity analysis over the parameter range described in Supplementary Table 8. Fig. 2 (**A**), (**B**) and (**C**) illustrate the impact of factors on the rate of spread of the vaccine-resistant genotype, assuming that vaccines were administered without a drug, and that the coverage was constant across deployment rounds. Results for the impact of all factors for each deployment strategy and various assumptions about coverage across rounds, and the use of drugs with the vaccine are similar and are shown in Supplementary Fig. 7-20.

Vaccine properties also play an important role. For an AIV, the second key driver is its initial efficacy (Fig. 2A). A higher initial efficacy increases the rate of spread as the selective advantage for the resistant genotype increases when the vaccine is more efficient (Fig. 2B). Moreover, the vaccine half-life also exerts a great influence in all cases (Fig. 2A). Longer half-lives lead to a higher rate of spread, as this extends the duration of selection of resistant parasites within hosts (Fig.2B).

The impact of entomological inoculation rate (EIR) and access to treatment (percentage of symptomatic cases who received treatment within 14-days of symptom onset) on the spread of resistance depends on the targeted age groups. When AIVs and BSVs are deployed to children, higher levels of EIR and lower levels of access to treatment increase resistance spread (Fig. 2B). In contrast, for vaccines targeting adults, these factors have opposite effects (Fig. 2C). For vaccines targeting the entire population, these factors have negligible effects (Fig. 2A). These findings are likely because, in high transmission settings, infections detectable by microscopy mainly occur in children. In contrast, in low transmission settings, infections mainly occur in adults (Supplementary Fig. 21). Therefore, for vaccines targeting children, the sensitive genotype has more opportunities to escape in the unvaccinated population (adults) at lower EIR compared to higher EIR, reducing the spread of resistance at lower EIR. The effect is reversed for vaccines targeting adults. Additionally, higher access to treatment reduces the proportion of infections among children more than among adults (Supplementary Fig. 21). Consequently, when adults are vaccinated, higher access to treatment reduces the opportunities for the sensitive genotype to escape in the unvaccinated population (children), favouring the spread of resistance. The effect is the opposite for vaccine-targeting children.

### Resistance dominance and loss of effectiveness

For each vaccine type and implementation strategy, we assessed how the degree of resistance impacted (i) the time required for the resistant genotype to spread from 1% to 50% frequency of inoculations, *T_50_* (Fig. 3A) and (ii) the vaccine effectiveness (VE) against clinical malaria assuming the resistant genotype reached 50% frequency (Fig. 3B) (see methods and Fig. 3 legends).

**Figure 3:**
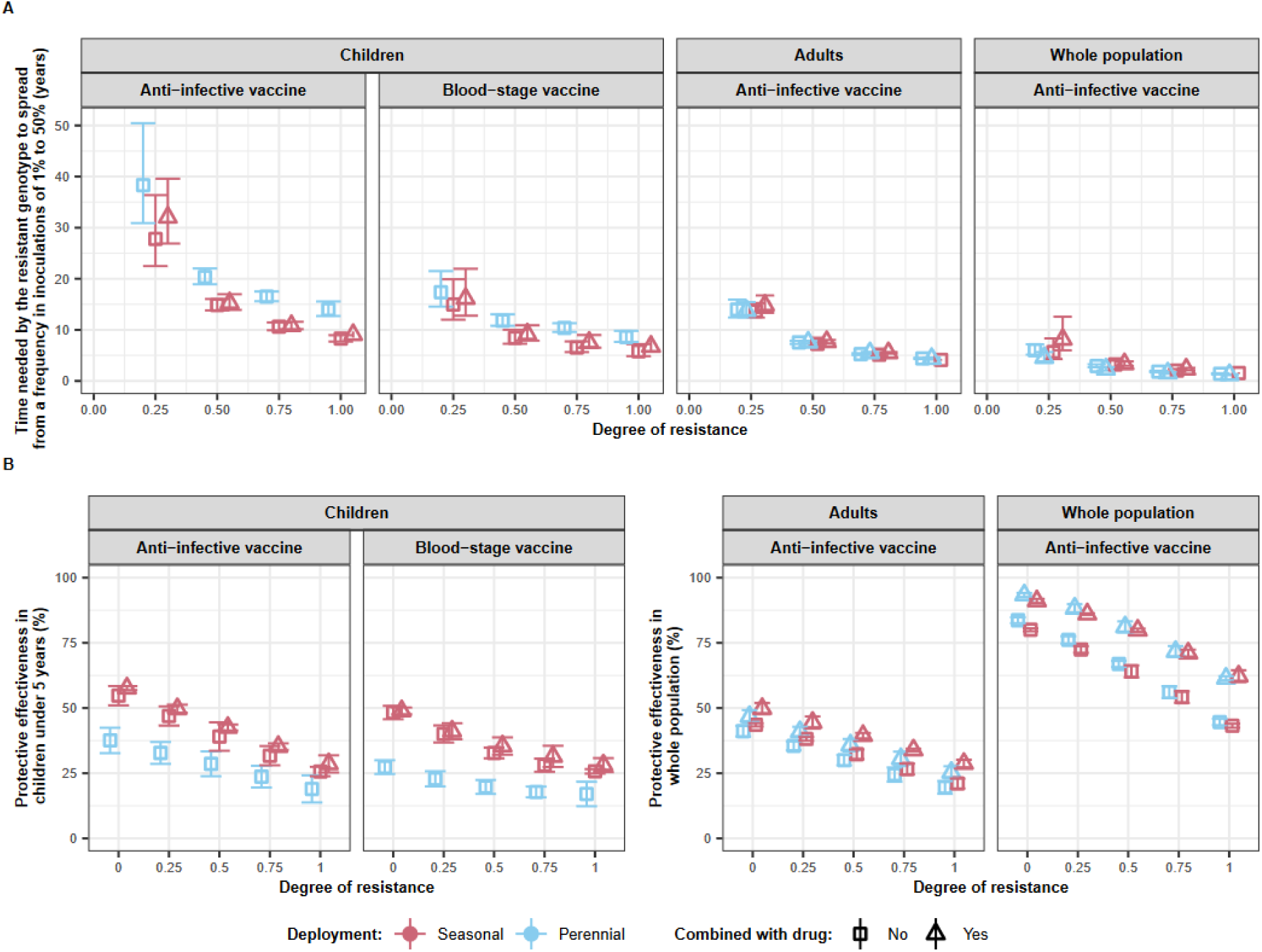
The impact of the degree of resistance on time needed before the resistant genotypes become dominant and vaccine effectiveness. (**A**) The predicted time needed for genotypes with different degrees of resistance (0, 0.25, 0.5, 0.75, 1) to vaccines to spread from a frequency in inoculations of 1% to 50%, *T_50_*. We predicted the *T_50_* and 95% confidence intervals for each vaccine type and deployment strategy (perennial deployment (light red marker), seasonal deployment with (light blue marker), with (triangle), or without (square) a drug implemented with the vaccines. (**B**) Estimated vaccine effectiveness (VE) and 95% confidence intervals in a population composed of 50% of sensitive genotypes and 50% of resistant genotypes with different degrees of resistance (0, 0.25, 0.5, 0.75, 1) to the vaccine. For vaccines deployed to children, the VE was estimated as the relative reduction in the number of clinical malaria cases in children under five years. For the AIV deployed to the whole population or adults, the VE was estimated as the relative reduction in the number of clinical malaria cases in the whole population. The VE was predicted for the same deployment strategies as above. For (**A**) and (**B**), we assumed that the vaccine had an initial efficacy of 95%, a half-life of 1 year, and was deployed with a coverage of 90% that did not decrease at the booster doses in a setting with an EIR of 5 inoculations per person per year, and low levels of access to treatment (25%). Results for different half-lives, transmission intensity, level of access to treatment, and assuming a final frequency of resistant genotype of 100% follow similar trends and can be seen in Supplementary Fig. 22-26.

The *T_50_* decreases dramatically with the degree of resistance (Fig. 3A), but even at a low degree of resistance, the resistant genotype becomes dominant relatively quickly. For example, when an AIV is deployed to children in a seasonal setting, the *T_50_* is estimated as 27.79 years (95% CI 22.48–36.38). This time reduces by 12.8 years for the BSV, suggesting that the spread may be faster for BSVs than AIVs. For vaccine strategies aiming at reducing transmission, the time until the vaccine-resistant genotypes becomes dominant is even shorter, as more individuals are vaccinated the more a resistant genotype is selected (Fig. 3A).

Nevertheless, the VE decreases almost linearly with increasing degrees of resistance. Thus, a vaccine is likely to provide high VE against genotypes with low degrees of resistance, but VE strongly reduces against genotypes with higher degrees of resistance. For example, the deployment of an AIV in a seasonal transmission setting leads to a VE of 54.8% (95% CI 51.8-57.7) when no parasites are resistant to the vaccine (degree of resistance of 0). The VE is reduced to 47.0% (95% CI 44.0-49.9) and 25.6% (95% CI 27.0-24.2) when the resistant genotype reaches a frequency of 50% and exhibits low (degree of resistance of 0.25) or full (degree of resistance of 1) resistance to the vaccine, respectively.

Furthermore, the *T_50_* is lower for seasonal deployments than for perennial deployments when targeting children (Fig. 3A). This outcome is due to the more frequent deployment of the vaccine in seasonal settings (Fig. 1B), leading to a higher VE (Fig. 3B) and selection pressure. However, when a vaccine is deployed to adults, or to the entire population, the deployment frequency is consistent in both seasonal and perennial settings, thus minimising differences in VE and *T_50_*.

Moreover, giving a drug alongside the vaccine increases the *T_50_* and the VE in most conditions (Fig. 3). The increase in *T_50_*is likely because the drug kills resistant genotypes that successfully infect individuals, thereby reducing spread. However, the impact is limited due to the drug’s shorter prophylactic period (around 20 days) compared to the vaccine (half-life of 1 year).

### The impact of combining vaccines

When the resistant parasite has a low degree of vaccine resistance, deploying an AIV to the entire population in a seasonal setting with a low transmission intensity may result in malaria elimination (Supplementary Fig. 27). We examined how a higher degree of resistance to the AIV could prevent malaria elimination, and if deploying the AIV with a TBV could mitigate the spread of resistance to the AIV sufficiently to enable elimination (see methods).

If an AIV is deployed without a TBV, the spread of resistant genotypes accelerates with the degree of resistance, leading to higher prevalence estimates 15 years after vaccine implementation (Fig. 4 when half-life TBV = 0 and initial efficacy of TBV = 0). Combining an AIV with a TBV can efficiently prevent the spread of genotypes resistant to the AIV and support reaching elimination. The spread of the resistant genotype to the AIV decreases more when the TBV has a longer half-life and higher initial efficacy (Fig. 4). With a half-life of 5 years (much longer than half-lives of current vaccines), the spread of genotypes fully resistant to the AIV is prevented, leading to the elimination of *P. falciparum* in all scenarios (Fig. 4).

**Figure 4:**
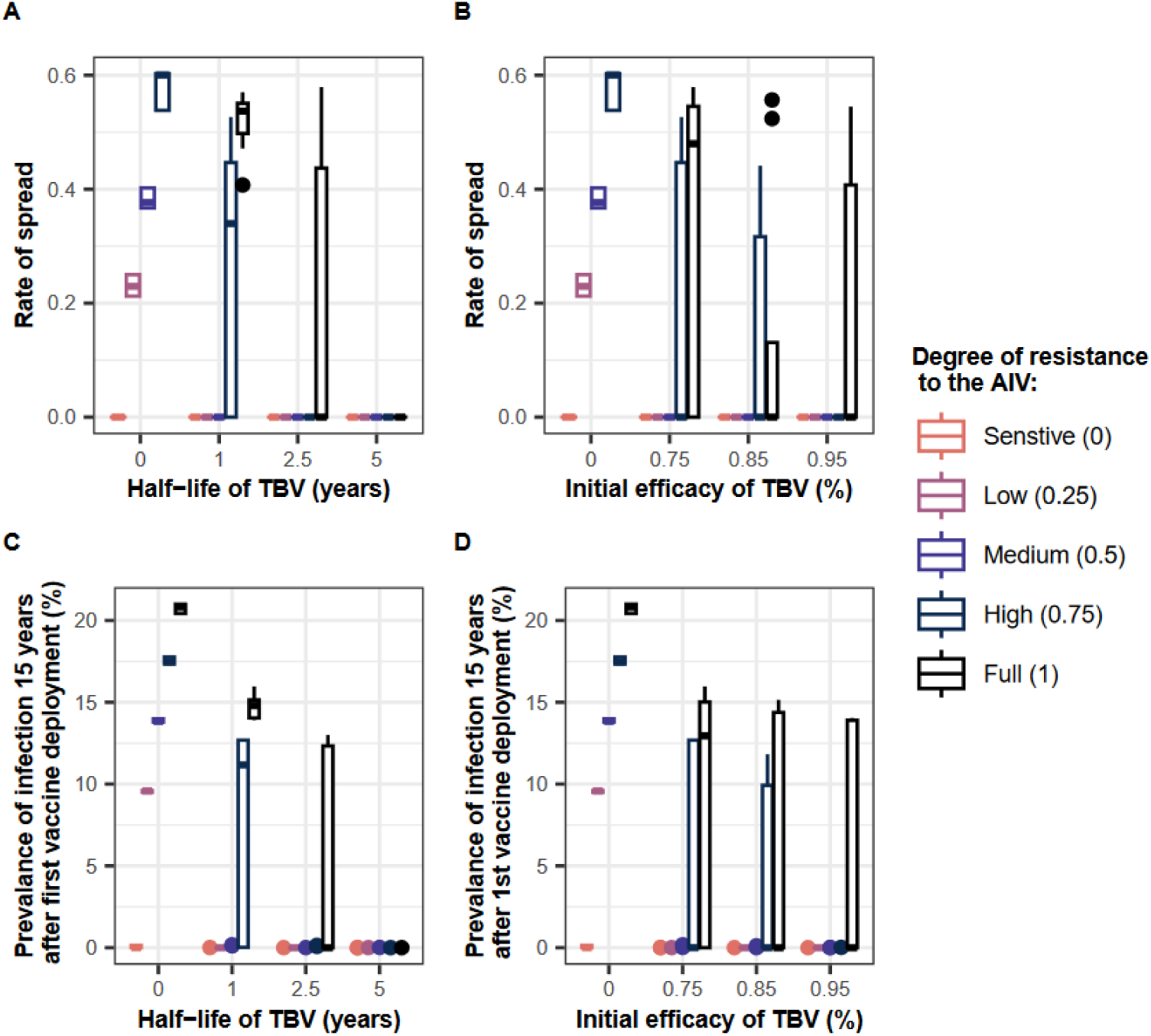
Impact of combining a TBV with an AIV. The impact of (**A**) the TBV half-life and (**B**) the TBV initial efficacy on the rate of spread of genotypes (the selection coefficient) with different degrees of resistance to the AIV (0 (orange boxplot), 0.25 (purple boxplot), 0.5 (blue boxplot), 0.75 (dark blue boxplot), and 1 (black boxplot)) when both vaccines were deployed together to the whole population. The impact of (**C**) the TBV half-life and (**D**) the TBV initial efficacy on the prevalence of malaria in the population 15 years after the implementation of the vaccines to the whole population infected with parasites having different degrees of resistance to the AIV. For (**A**), (**B**), (**C**), and (**D**), we evaluated the outcomes for different combinations of the TBV half-life (1, 2.5, 5 years) and the TBV initial efficacies (0.75%, 0.85%, 0.95%). The AIV and TBV were deployed in combination to the whole population every two years at a coverage of 90%. The AIV had a half-life of 2 years and an initial efficacy of 95%. The vaccines were deployed in a seasonal setting with low transmission intensity (10 inoculations per person per year) and high access to treatment (60%). The vaccines were deployed without drugs, assuming the coverage was constant at every round.

## Discussion

Previous studies have highlighted that antigens of malaria vaccines exist in multiple alleles^1–3^, probably due to acquired immunity that favour their diversification. Although it remains uncertain whether this diversity is associated with variation of efficacy for some vaccines, we show that if vaccine-resistant genotypes exist, they could spread rapidly and strongly reduce the VE of vaccines depending on their degree of resistance (defined as the relative decrease in the initial vaccine efficacy of the resistant genotype compared with the sensitive one). While malaria vaccines will save lives and their deployment should not be delayed, these results call for a comprehensive assessment of how VE may vary between genotypes. This would involve both an assessment of the existence of vaccine-resistant genotypes against current malaria vaccines and those under development in areas where we plan to implement vaccines and also a framework to assess the difference in vaccine efficacy between genotypes. Lastly, we should be prepared to assess strategies to develop and implement vaccines that minimise the risk of spread if the risk of resistance is assessed to be high.

The rapid spread of genotypes with a low degree of resistance to vaccines will have a minor effect on the VE, whereas highly resistant genotypes can spread even faster and diminish the VE more strongly. Therefore, it is crucial to better understand the diversity of targets of current vaccines and those under development and assess how this diversity is associated with differences in efficacy. This will involve defining how to measure the degree of resistance of resistant genotypes using studies and developing assays discussed in Box 1. Once the diversity in antigens and the difference in efficacy between genotypes is better understood, vaccine developers should prioritise targeting conserved antigens (with only one or few alleles), such as RH5^45^, Pfs25^46,47^ and Pfs230^47^, or include all alleles that are immunologically relevant in the formulation. However, as these antigens may not be constrained, the parasites can still accumulate mutations that lead to vaccine resistance. Therefore, it is essential to implement molecular surveillance and monitor vaccine efficacy to detect the potential emergence and spread of vaccine-resistant genotypes and react accordingly, as elaborated in Box 1.

We demonstrated that the vaccine properties and their mode of action on malaria (prevent infection, morbidity or transmission) have a crucial influence on the spread of resistance. Longer half-lives and higher initial efficacy favour the spread of vaccine resistance, as the advantage of the resistant genotype becomes greater when the vaccine is more efficient. Moreover, resistance spread is much faster for BSVs than AIVs. This is because an AIV can prevent resistant infection transmission when it successfully prevents the infection. However, the BSV can only partially reduce the multiplication of the resistant parasite at the blood-stage, which can potentially reduce gametocytes but may not entirely block transmission. Furthermore, if no resistant genotype pre-exists, the parasite population is much larger during the blood-stage than the liver-stage (Fig. 1A)^44^, increasing the likelihood of a mutation conferring vaccine resistance to emerge. Therefore, it may be more challenging to develop a highly efficient BSV (long half-life and high initial efficacy) that is evolution-proof compared to an AIV.

If vaccine-resistant genotypes are present in some regions, our results suggest that vaccine developers should consider combining vaccines that target different stages to reduce the spread of these genotypes, as discussed in Box 1. However, combining a drug with a vaccine only slightly reduces the spread of the resistant genotype as the drug offers a short prophylactic period compared to the vaccine. Further studies could explore if more frequent drug administration, such as during seasonal malaria chemoprevention, could further reduce the resistance spread.

Moreover, our results suggest the rate of spread accelerates when a larger range of age groups are vaccinated. Vaccines that aim to achieve elimination will likely need to target a large proportion of the population, which will impose a strong selection pressure. An alternative solution could be to implement different vaccines for children and adults, thereby reducing the number of individuals selecting the same genotype and creating a heterogeneity of selection.

As with all models, we cannot capture all the complex aspects of malaria and the uncertainties surrounding parasite, vaccines, immunity, resistance etc. First, we did not model the recombination of the *P. falciparum* genome in mosquitoes. If various mutations in different genes are required to confer resistance, recombination could reduce the spread of resistance by separating these mutations when they are present at low frequencies^48^. However, recombination is unlikely to impact the spread of resistance to vaccines that target a single protein (subunit vaccines), but could reduce the spread of resistance to combination vaccines and whole sporozoite vaccines. Second, we modelled only two genotypes in order to extract key conclusions. In reality, there are probably many more genotypes which lead to increased competition that would reduce the spread of the most resistant genotype. However, before including more genotypes in mathematical models, it is crucial to evaluate the extent of antigen diversity that is immunologically relevant. Third, we did not model *denovo* mutations and assumed that the resistant genotype exists in the population. While this assumption may hold for AIVs, where the pre-erythrocytic cycle is too short for within-host mutations, this may not hold for the blood-stage (Fig. 1A). If we investigated the emergence of mutations, the time until the resistant genotype is dominant would be prolonged by the time required for the mutation to occur and establish (see more previous work^49,50^). Finally, we assumed that there were no fitness costs associated with the resistant genotype, as our focus was on a genotype that existed without selection pressure imposed by vaccines. If we explored varying fitness costs, the relationships between the factors presented here would remain. However, the rate of spread would likely be lower, and thus the time until the resistant genotype would be dominant would be longer.

In conclusion, the potential rapid spread of vaccine-resistant genotypes and their potential impact on vaccine effectiveness calls for a deeper understanding of whether existing genotypes exhibit reduced sensitivity to current and future malaria vaccines. Our study reports on strategies to identify and reduce the risk and spread of vaccine resistance ( box1). While this is the first quantitative study to systematically explore the influence of many factors on the risk of selection from malaria vaccines, clinical studies have shown selection of resistance against BSVs^14^. Further studies should confirm or disprove this result for AIVs by investigating whether implementing vaccines, such as RTS,S/AS01 or R21, leads to the spread of vaccine-resistant genotypes (genomic monitoring is already ongoing for these vaccines and other therapeutics) and how it impacts the vaccine’s efficacy.

#### Box 1: List of recommendations to evaluate and reduce the risk of resistance to current and future malaria vaccines

1. **We need to better understand if genotypes with reduced efficacy to current and future vaccines exist. This will involve:**

1.1 Population genomic studies to assess the cross-sectional diversity in the antigen targets of current and future malaria vaccines in geographies where they are likely to be deployed, as well as longitudinal studies to understand how antigenic diversity changes over time in the absence of vaccine pressure.
1.2 If antigen targets are not conserved, genotypically variable efficacy should be assessed in all phase 2/3 trials, and assays should be performed to assess if different alleles are associated with variations in vaccine efficacy. This could involve preclinical assays such as blood-stage growth inhibition assays, liver-stage development assays, and membrane-feeding assays performed with different genotypes. These assays will likely require development efforts to ensure they are sufficiently sensitive to variables affecting in vivo infections. Ideally, such assays would be performed at early stages of development to identify the risk of resistance and prioritise the development of vaccines with lower risk or include the immunologically relevant alleles of the antigen in vaccines.
2. **If the risk of resistance is high for some vaccines, or if the deployment may impose significant selection pressure, strategies must be considered to delay the spread, such as:**

2.1 Combining multiple vaccines that target different antigens. If combination vaccines are developed, the establishment and spread of a genotype with a mutation in one of the antigens will be delayed by the other antigens that remain effective on the genotype. Ideally, the targeted antigens should be conserved or include the different immunologically relevant alleles to ensure that no resistant genotype exists.
2.2 Implementing multiple vaccines in the same population. Once multiple vaccines are approved, health ministries could implement multiple vaccines in the same population to create heterogeneity of selection and potentially reduce the spread of vaccine resistance. This will be particularly important for vaccines that aim to achieve elimination, as they will likely need to target a large proportion of the population. Thus, different vaccines could be deployed to different population groups, such as children and adults.
3. **Once vaccines are implemented, genotype diversity and vaccine effectiveness should be monitored and vaccination programs must take action if vaccine effectiveness is impacted. This involves:**

3.1 Implementing molecular surveillance of vaccine antigens to monitor change in antigen diversity and regularly
3.2 vaccine effectiveness, as done for malaria drug resistance. In vitro studies could be utilised to assess the association between the new allele and vaccine efficacy using similar assays discussed in point 1.2.
3.3 Developing a platform to rapidly adapt vaccine formulation to include new alleles if appropriate. If a new genotype strongly undermines the vaccine’s effectiveness, the vaccine could be modified to incorporate the new allele in its formulation and restore the vaccine’s efficacy. This approach was previously employed mRNA vaccines during the SARS-CoV-2 pandemic. However, adapting subunit vaccines or live-attenuated vaccines, especially for parasites, may require much more time.

## Methods

### 2.1. Model description and parameterizations

We used our existing individual-based model of malaria transmission, OpenMalaria, that simulates *P. falciparum* transmission and epidemiology in mosquitoes and humans. The model has been described in detail in previous work^51–53^ and is described in Supplementary Note 1.1. Our model explicitly captures the mechanisms of immunity acquisition and was previously used to predict the potential public health impact of AIVs, BSVs, and TBVs^54^. We previously adapted the model to track multiple parasite genotypes with different sensitivities to drugs and to model parasite within-host dynamics explicitly^49^. Here, we further modified the model to capture that genotypes can have different degrees of resistance to vaccines. In our model we assumed that each modelled vaccine has a specific initial efficacy which decays over time biphasically with a specified half-life (Supplementary Note 1.1). We simulated a range of efficacies, and half-lives during the global sensitivity analysis (see methods section 2.2 and Supplementary Table 8). For a resistant genotype, we assumed the initial efficacy is reduced compared to the sensitive genotype. We defined the degree of resistance of the parasites to the vaccine as the relative decrease in the initial vaccine efficacy of the resistant genotype compared with the sensitive one. We simulated a range of degrees of resistance during the global sensitivity analysis (see methods section 2.2 and Supplementary Table 8). In our model, an AIV prevents a proportion of infections from reaching the blood-stage (Fig. 1A). A BSV reduces the multiplication rate of the parasite within-host (Fig. 1A). A TBV diminishes the probability of parasite transmission from humans to mosquitoes (Fig. 1A).

Following the recommendations from WHO for RTS,S/AS01^7^, we simulated the deployment of AIVs and BSVs in children to prevent morbidity using two different deployment strategies that depend on the seasonality of the setting. In perennial transmission settings, the primary vaccination series was delivered to children at nine months of age with a booster dose 18 months later (Fig. 1B). We assumed transmission occurs during four months in seasonal settings and we delivered a primary vaccination series to children at nine months of age via the Expanded Program of Immunization, and children older than 15 months and less than five years of age received a booster dose two months before the transmission peak in four consecutive years (Fig. 1B). In seasonal settings, we additionally assessed the spread of vaccine resistance when a drug frequently used for chemoprevention is deployed with each booster (sulfadoxine-pyrimethamine + amodiaquine, see parameterisation in Supplementary Note 1.2)^55^). This allowed us to assess whether deploying the vaccine with a drug can affect the spread of resistance to a vaccine.

To prevent transmission, we explored scenarios when an AIV was deployed either to the whole population or only adults. We simulated vaccine deployment to the targeted people through mass administration with a booster dose every two years in both seasonal and perennial settings (Fig. 1C). In the seasonal setting, mass administration occurred two months before the peak of transmission (Fig. 1C). For each setting, we also assessed the spread of vaccine resistance both with and without a drug combination frequently used from mass drug administration at the same time as the vaccine (dihydroartemisinin–piperaquine, see parameterisation in Supplementary Note 1.2).

For each use case, we simulated a range of deployment coverages during the global sensitivity analysis (see section 2.2 and Supplementary Table 8) and assumed booster vaccine coverage was either the same or reduced by 20% compared to the initial deployment coverage. We assumed that the efficacy conferred by each vaccine dose was the same for the primary vaccination series as for each booster dose. During the global sensitivity analysis, we simulated a range of settings that varied in annual transmission intensity and level of access to treatment (see section 2.2 and Supplementary Table 8).

### 2.2. Assess the drivers of resistance spread

For each seasonality setting, deployment strategy and vaccine type, we systematically quantified the influence of multiple epidemiological, vaccine, health system, and deployment factors on the spread of vaccine-resistant genotypes using global sensitivity analyses (Fig. 1D). Global sensitivity analysis allows us to estimate how strongly variation in the above-mentioned factors impact the rate of spread of the resistant genotype. This method required first defining a range of interest for each factor (such as vaccine properties, coverage, transmission intensity, access to treatment, and degree of resistance) as specified in Supplementary Table 8. We then simulated and estimated the rate of spread of the resistant genotype in a population of sensitive parasites for a set of random combinations of factor values. The rate spread of the resistant genotype was assessed through the selection coefficient, defined as the rate at which the logit of the resistant genotype frequency increases each parasite generation (Supplementary Note 2.1)^49,50^.

However, the global sensitivity analysis requires simulating many (200,000) random combinations of factors values. Thus, to be more efficient, we used a workflow previously developed to assess the key drivers of the spread of drug resistance (Supplementary Note 2.1)^49^. This workflow involved to trained an emulator to learn the relation between input parameters and the rate of spread from a smaller amount (2500) of model simulations. The fit of the emulator could be improved by adaptative sampling. Once the emulator reached a good fit, the emulator was used to predict the rate of spread for a large amount of combination of factors values and perform the global sensitivity analysis (Supplementary Note 2.1)^49^. The global sensitivity analysis was performed using Sobol’s method of variance decomposition (Supplementary Note 2.1)^56^, which allowed us to estimate first-order indices of each factor (representing their influence on the rate of spread) and the 25th, 50th, and 75th quantiles of the predicted rate of spread across each parameter range.

### 2.3. Estimate time to dominance of resistance

To estimate the impact of vaccine type and deployment strategy in a timeframe, we used the trained emulators to predict the selection coefficient for a set of parameter combinations and translated the selection coefficients to the time needed for the resistant genotype to spread from 1% to 50% frequency of inoculations, *T_50,_* as described in Supplementary Note 2.2^49^. Note that in the real world, the resistant genotype could exist at a higher frequency than 1% leading to a shorter time to reach 50% frequency. We predicted the *T_50_* for each vaccine type, deployment strategy, and seasonality setting when AIVs or BSVs were deployed to children to reduce morbidity or when the AIV was deployed to adults or the whole population to reduce malaria transmission. In each case, we modelled a vaccine with an initial efficacy of 95% and a half-life of 1 or 1.5 years. We assumed the resistant genotype had various degrees of resistance to the vaccine (0, 0.25, 0.5, 0.75, and 1). The vaccines were deployed at a coverage of 90% to illustrate high selection pressure in settings with different transmission intensities (5 or 50 inoculations per person per year) and levels of access to treatment (25 or 60%).

### 2.4. Estimate vaccine effectiveness

For each vaccine type and deployment strategy, we estimated the impact of the spread of vaccine-resistant genotype on the protective effectiveness (PE) of the vaccine. To do so, we evaluated the PE against parasite populations containing 50% of sensitive genotypes and 50% of resistant genotypes for which the vaccine had different degrees of resistance (0, 0.25, 0.5, 0.75, and 1). For vaccines that reduce morbidity, we defined the PE as the relative reduction of incidence of clinical malaria cases in children under five years during the year following the time at which the first cohort of vaccinated children received their last vaccine booster doses compared to the incidence of clinical malaria the year before the vaccine implementation (Supplementary Note 2.3 and Supplementary Fig. 5). For vaccines that reduce malaria transmission, we assessed the PE as the relative reduction of incidence of clinical malaria cases in the whole population during the two years following vaccine implementation compared to the incidence of clinical malaria cases during the two years before vaccine implementation (Supplementary Note 2.3 and Supplementary Fig. 6). We estimated the PE for the same combination of parameters described in the section above.

### 2.5. The impact of combining vaccines

We further explored how deploying a TBV with an AIV to the whole population can delay the spread of genotype resistant to an AIV. To do so, we first selected simulations where the implementation of an AIV to the whole population led to the elimination of *P. falciparum* (defined as the prevalence of infection in the entire population below 0.1%) in the seasonal setting (Supplementary Fig. 27). We did not consider the perennial setting as elimination in our simulations was less frequent in this setting. In the selected simulation, the AIV was deployed at a coverage of 90% to the whole population in a setting with low transmission intensity (10 inoculations per person per year) and high access to treatment (60%). The AIV had a half-life of 2 years and an initial efficacy of 95%. Then, we estimated how different degrees of resistance to the AIV (0, 0.25, 0.5, 0.75, 1) affected the spread of resistance to the AIV and the prevalence of infection in the whole population 15 years after vaccine deployment. Note that when *P*. *falciparum* elimination occurred, we set the selection coefficient to zero. Finally, we assessed how combining a TBV with an AIV could reduce the spread of the genotype resistant to the AIV, and the prevalence of infection in the whole population 15 years after vaccine deployment. We assumed that the resistant genotype was fully sensitive to the TBV and modelled that the TBV had various initial efficacies (75, 85 and 95%) and half-lives (1, 2.5 and 5 years).

## Data, code, and materials availability

All data and code used to produce the figures are available at https://doi.org/10.5281/zenodo.10418576. OpenMalaria code version 44.0 (https://github.com/SwissTPH/openmalaria) and documentation (https://github.com/SwissTPH/openmalaria/wiki) are open access and open source. The code used to run the simulations and perform the analyses can be found at https://doi.org/10.5281/zenodo.10418569.

## Acknowledgements

This study was funded through the Swiss National Science Foundation Professorship grant of M.A.P. (PP00P3_203450). Calculations were performed at sciCORE (http://scicore.unibas.ch/) scientific computing centre at the University of Basel.

## Author contributions

M.A.P. conceived the study. T.M., T.L., J.M., and M.A.P. designed the study. A.C. modified the model OpenMalaria and code. T.M. performed the analyses. T.M., T.L., J.M., S.L.K., and M.A.P. interpreted the results and examined their implications. T.M. wrote the first draft of the manuscript and created the tables and figures. T.L., S.L.K., J.M., A.C. and M.A.P. revised the manuscript. All authors reviewed and approved the final manuscript. M.A.P. acquired the funds and managed the project.

## Additional information

**Supplementary Information** is available for this paper.

**Correspondence and requests** for materials should be addressed to M.A.P.

**Reprints and permissions** information is available at www.nature.com/reprints.

## Supplementary Information

### 1 Supplementary Note 1: Malaria transmission model and vaccine parameterisation

#### 1.1 OpenMalaria

We adapted our individual-based model of malaria transmission and epidemiology, OpenMalaria^1–3^, to track the spread of a genotype with reduced sensitivity to vaccines. OpenMalaria explicitly models mechanisms of immunity acquisition and has previously been used to predict the public health impact of anti-infective vaccines (AIVs), blood-stage vaccines (BSVs), and transmission-blocking vaccines (TBVs)^4^. Our model was also formerly adapted to track multiple genotypes with different drug sensitivities and mechanistically model parasites within-host dynamics^5^. We further adjusted our model to capture that genotypes can have varying degrees of resistance to the vaccine. Our model has been described in detail in previous work^1–3,5,6^. Here, we briefly describe the main attributes of our model, with a focus on the novelty that allowed us to model vaccine resistance.

OpenMalaria is an individual-based model, meaning that each individual is tracked separately throughout their life. OpenMalaria runs on a five-day time step and consists of an ensemble of models that simulate the epidemiology and transmission of malaria in mosquito vectors and human hosts^1–3^. The vector model of OpenMalaria is a periodically forced deterministic model that captures the life and feeding cycle of mosquitoes^1^. During the feeding cycle, the model tracks the infectious states of the mosquitoes^1^. In addition, the model can simulate seasonality thanks to its periodicity^1^. The number of newly infected human hosts depends on the simulated entomological inoculation rate (EIR)^1^.

The human component of OpenMalaria simulates infections within humans who are bitten by an infected mosquito. Liver-stage immunity in humans reduces the number of infections reaching the blood-stage based on their previous exposure to the parasite^3^. The parasite dynamics at the blood-stage are simulated using a mechanistic within-host model that calculates the parasite densities based on the parasite multiplication rate and the effects of innate, acquired variant-specific, and acquired variant-transcending immunities^7^. Individuals develop their immunity based on their cumulative exposure to the parasite^8^. Infants are protected by maternal immunity during the first months of life^8^. The simulated parasite densities influence diagnostic outcomes, patient symptoms^9^, risks of severe malaria and death^10,11^, and the probability that a feeding mosquito becomes infected^12^.

Patients with uncomplicated malaria and severe malaria receive treatment as described by a case management model^3^. In our model we can specify the diagnostic sensitivity and specificity, the rate of treatment access, and the treatment provided to symptomatic patients. The drug concentration in the patient is described using a pharmacokinetic (PK) model (with one-, two- or three-compartments depending on the PK dynamics of the drug)^5,13^. The effect of drug concentration on the within-host parasite multiplication rate is estimated using a pharmacodynamic (PD) model^5,13^.

In addition, our model allows the deployment of various interventions, including vaccines that target different life cycle stages^4^. OpenMalaria models that an AIV prevents a proportion of infections from reaching the blood-stage (efficacy of 50% means that 50% of infections do not reach the blood-stage). The model assumes that a BSV reduces the multiplication rate of the parasite within-host (efficacy of 50% implies that the multiplication rate of the parasite is reduced by half)^4^. OpenMalaria captures that a TBV diminishes the probability of transmission from humans to mosquitoes (efficacy of 50% means that the vaccine halved the transmission probability)^4^. For each vaccine type, the user needs to define a specific initial efficacy that decays over time, assuming a specific half-life and a decay function. Here we assumed that the decay function followed a Weibull function with a decay shape parameter of 0.69 to capture a bi-phasic (fast decay followed by slow decay) decay. This decay function captures the decay of the RTS,S/AS01 vaccine^14^. In OpenMalaria, the vaccine can be deployed to individuals reaching a specific age or deployed at a defined time to individuals within a particular age range.

Recently, our model was adapted to specify multiple parasite genotypes with varying degrees of drug sensitivity^5^. With this update, the pharmacodynamics parameters can be parameterised individually for each genotype to model different degrees of drug susceptibility^5^. Moreover, our model allows for concurrent infection of multiple parasite genotypes within the same host and captures that different parasite genotypes indirectly compete as immunity regulates the total parasite load^5^. Users can also specify a fitness cost associated with mutations that confer drug resistance by reducing the within-host multiplication rates for the resistant genotype^5^. When a human is infected, the genotype of the parasite is drawn from genotype frequencies in human infections from the previous five-time steps^5^, which allows us to model the spread of the genotype in the population.

Here, we further adapted our model to capture that genotypes can have varying degrees of vaccine sensitivity. We refer here to the degree of resistance as the relative reduction of the initial efficacy for the resistant genotype compared to the sensitive one.

#### 1.2 Parameterisations of chemopreventive drugs deployed with the vaccines

We investigated the spread of vaccine-resistant genotypes for the two future use cases of malaria vaccines: 1) vaccines to reduce morbidity and mortality in children and 2) vaccines aiming to reduce transmission in the entire population.

For the first use-case we focused on the spread of genotypes resistant to vaccines with several modes of action implemented to prevent morbidity and mortality in children. The vaccines could be either an AIV or a BSV. The AIV and BSV were administered in children living in a setting with seasonal (with transmission occurring mainly over four months) or perennial transmission as described in the methods of the paper and Fig. 1B. In the seasonal setting, we assessed the spread of vaccine resistance when deploying a drug combination with or without each vaccine booster. We assumed that the drug combination used was sulfadoxine-pyrimethamine+amodiaquine, which is commonly used for seasonal malaria chemoprevention in children^15^. We assumed the same dosage and schedule as sulfadoxine-pyrimethamine (SP) for seasonal malaria chemoprevention (Supplementary Table 1)^15^. We modelled amodiaquine (AQ) and its active metabolite desethylamodiaquine (DEAQ) with two two-compartment PK/PD models with first-order absorption as in previous work^16^ (Supplementary Tables 2 and 3). We modelled SP as a single, long-acting drug with one-compartment PK/PD and first-order absorption as in previous work^16^ (Supplementary Table 4).

For the second use case, we focused on the spread of genotypes resistant to an AIV vaccine that would be implemented in the entire population or in adults to reduce malaria transmission. The AIV was deployed in seasonal and perennial transmission settings, as detailed in the methods section and Fig. 1C. For each setting, we assessed the spread of vaccine resistance when deploying the vaccine alone or with a drug combination. The drug chosen for this scenario was dihydroartemisinin–piperaquine, which is commonly used for mass drug administration. The dosage and schedule employed are provided in Supplementary Table 5. Dihydroartemisinin was modelled with one-compartment PK assuming instantaneous absorption (Supplementary Table 6) as in previous work^17,18^. Piperaquine was modelled using a two-compartment PK/PD model with first-order absorption as in our previous work^19^ (Supplementary Table 7).

**Supplementary Table 1:** Dosage regimens of sulfadoxine/pyrimethamine and amodiaquine by age group.

| Age group | Sulfadoxine/pyrimethamine | Amodiaquine | Reference |
| --- | --- | --- | --- |
| 3 months to 1 year | Single dose of 250/12.5 mg | Three daily doses of 76.5 mg | *15* |
| 1 to 5 years | Single dose of 500/25 mg | Three daily doses of 153 mg | *15* |

**Supplementary Table 2:** Amodiaquine pharmacokinetic/pharmacodynamic parameters.

| Parameter | Value | Reference |
| --- | --- | --- |
| Absorption rate (per day) | 2.16 | <sup>16</sup> |
| Elimination rate (per day) | 42.00 | <sup>16</sup> |
| Rate at which the drugs move from the central compartment to the peripheral compartment (per day) | 51.00 | <sup>16</sup> |
| Rate at which the drugs move from the peripheral compartment to the central compartment (per day) | 1.46 | <sup>16</sup> |
| Volume of distribution (l/kg) | 8.00 | <sup>16</sup> |
| Maximum killing rate (per day) | 2.30 | <sup>16</sup> |
| EC50 (mg/l) | 0.0043 | <sup>16</sup> |
| Slope of the effect curve | 7.00 | <sup>16</sup> |

EC50=half-maximal effective concentration.

**Supplementary Table 3:** Desethylamodiaquine pharmacokinetic/pharmacodynamic parameters.

| Parameter | Value | Reference |
| --- | --- | --- |
| Absorption rate (per day) | 2.16 | <sup>16</sup> |
| Elimination rate (per day) | 0.86 | <sup>16</sup> |
| Rate at which the drugs move from the central compartment to the peripheral compartment (per day) | 1.70 | <sup>16</sup> |
| Rate at which the drugs move from the peripheral compartment to the central compartment (per day) | 0.46 | <sup>16</sup> |
| Volume of distribution (l/kg) | 18.40 | <sup>16</sup> |
| Maximum killing rate (per day) | 2.30 | <sup>16</sup> |
| EC50 (mg/l) | 0.02 | <sup>16</sup> |
| Slope of the effect curve | 7.00 | <sup>16</sup> |

EC50=half-maximal effective concentration.

**Supplementary Table 4:** Pharmacokinetic/pharmacodynamic parameters for sulfadoxine/pyrimethamine.

| Parameter | Value | Reference |
| --- | --- | --- |
| Volume of distribution (l/kg) | 0.29 | <sup>16</sup> |
| Absorption rate (per day) | 12.50 | <sup>16</sup> |
| Elimination rate (per day) | 0.12 | <sup>16</sup> |
| Maximum killing rate (per day) | 2.30 | <sup>16</sup> |
| EC50 (mg/l) | 2.40 | <sup>16</sup> |
| Slope of the effect curve | 2.10 | <sup>16</sup> |

EC50=half-maximal effective concentration.

**Supplementary Table 5:** Dosage regimens of dihydroartemisinin–piperaquine per kilo.

| **Drug** | **Dosage per kilo** |
| --- | --- |
| Dihydroartemisinin | Three daily doses of 4 mg/kg |
| Piperaquine | Three daily doses of 20 mg/kg |

**Supplementary Table 6:**
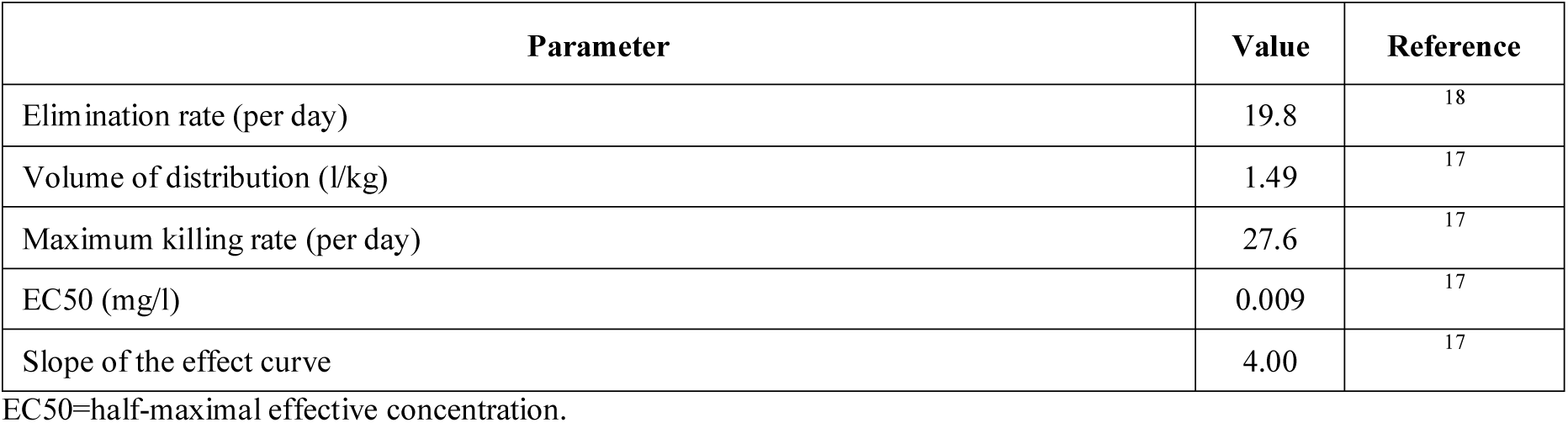
Dihydroartemisinin pharmacokinetic/pharmacodynamic parameters.

| Parameter | Value | Reference |
| --- | --- | --- |
| Elimination rate (per day) | 19.8 | <sup>18</sup> |
| Volume of distribution (l/kg) | 1.49 | <sup>17</sup> |
| Maximum killing rate (per day) | 27.6 | <sup>17</sup> |
| EC50 (mg/l) | 0.009 | <sup>17</sup> |
| Slope of the effect curve | 4.00 | <sup>17</sup> |

EC50=half-maximal effective concentration.

**Supplementary Table 7:** Piperaquine pharmacokinetic/pharmacodynamic parameters.

| Parameter | Value | Reference |
| --- | --- | --- |
| Absorption rate (per day) | 11.16 | <sup>19</sup> |
| Elimination rate (per day) | 0.62 | <sup>19</sup> |
| Rate at which the drugs move from the central compartment to the peripheral compartment (per day) | 8.46 | <sup>19</sup> |
| Rate at which the drugs move from the peripheral compartment to the central compartment (per day) | 3.31 | <sup>19</sup> |
| Volume of distribution (l/kg) | 173.00 | <sup>19</sup> |
| Maximum killing rate (per day) | 3.45 | <sup>18</sup> |
| EC50 (mg/l) | 0.02 | <sup>18</sup> |
| Slope of the effect curve | 6.00 | <sup>18</sup> |

EC50=half-maximal effective concentration.

### 2 Supplementary Note 2: Additional methods

#### 2.1 Additional methods for exploring the rate of spread

For each seasonality setting, deployment strategy, and vaccine type, we systematically quantified the influence of multiple epidemiological, vaccine, health system, and deployment factors (Supplementary Table 8) on the spread of vaccine-resistant genotypes using global sensitivity analyses. However, each global sensitivity analysis required 200,000 simulations (see details below). It was unfeasible to simulate this amount of simulation with our model. To be computationally efficient, we trained an emulator on a set of OpenMalaria simulations (2500 simulations, see details below). Emulators are predictive models that can approximate the relationship between the input and output parameters of complex models and can run much faster than complex models^20^. The emulator fit to the data was improved through adaptive sampling. We then used fitted emulators to perform the global sensitivity analysis using Sobol’s method of variance decomposition^21^. We summarised the steps of our workflow below and in Fig. 1D and have previously detailed a similar workflow for drug resistance^16^.

##### 2.1.1 Sample combinations of parameters

We established the parameter range for each investigated factor to include feasible parameter values as follows (Supplementary Table 8). The range for vaccine coverage included values from unoptimistic to optimistic levels. The range for vaccine’s initial efficacy captured vaccines with low to high efficacy. The vaccine half-life range enclosed short half-lives, such as the RTS,S/AS01 vaccine (half-life of 7 months)^22^, and optimistic longer half-lives that could be provided by future vaccines. The half-life range included a longer half-life vaccine aimed to reduce transmission. The degree of resistance varied from almost fully sensitive to fully resistant, based on the findings of Neafsey *et al.*, which did not reject the hypothesis that the RTS,S/AS01 vaccine may be fully ineffective against genotypes with more than five mutations compared to the vaccine genotype^23^. The EIR ranged from low to high transmission intensities, as observed in regions where vaccines are/will be administered. In the case of uncomplicated malaria, vaccinated and unvaccinated individuals received an artemisinin-combination-based therapy (ACT), as recommended by the World Health Organization (WHO)^24^. The range for access to treatment captured settings with low to high access to treatment defined as the percentage of symptomatic cases who received treatment within 14-days of symptom onset. The ACT was equally efficient against the vaccine-sensitive and vaccine-resistant genotypes.

We employed a Latin hypercube sampling algorithm (LHS)^25^ to randomly sample 250 parameter combinations from the parameter spaces.

##### 2.1.2 Simulations and estimation of the spread

We simulated each parameter combination using five stochastic realisations. In each simulation, we tracked 100,000 individuals with an age distribution similar to that of Tanzania, a lower-middle-income country^26^. We started the simulations with a frequency of 50% for each of the two genotypes. We did not start with a lower frequency in order to mitigate the effects of genetic drift (defined as the change in frequency of an existing genotype in the population due to random chance), which would have increased the chance of stochastic extinction of the resistant genotype and reduced the accuracy of the estimated rate of spread^5,27^. Nevertheless, since the rate of spread was not frequency-dependent in our model, our estimates reflect selection occurring at a low frequency^5,27^.

The simulations started with a burn-in period of 130 years, after which the vaccine was deployed for 15 years. For vaccines that prevent morbidity, we started to estimate the rate of spread when the first cohort of vaccinated children received their final booster doses (Supplementary Fig. 1) and stopped ten years later. For vaccines that prevent transmission, we started to estimate the rate of spread after the first deployment of the vaccine and stopped ten years later (Supplementary Fig. 2). The rate of spread was assessed through the selection coefficient, which represents the rate at which the logit of the resistant genotype frequency increases each parasite generation^5,27^. The selection coefficient, *s*, was estimated as followed,

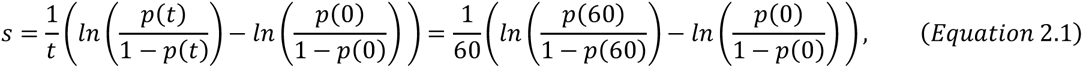

where *t* is the time in number of parasite generations assuming that a parasite generation was equal to 60 days^5,27^, and *p(t)* is the frequency of the resistant genotype at time *t*. The frequency of the resistant genotype was determined based on the frequency of each genotype in inoculations (the number of inoculations carrying the resistant genotype divided by the sum of the number of inoculations carrying the resistant genotype and the number of inoculations carrying the sensitive genotype).

##### 2.1.3 Model emulation and adaptive sampling

After estimating the rate of spread, we randomly divided our dataset into a training and a testing dataset. The training dataset was 80% of the simulations, while the test dataset was the remaining 20%. The training dataset was used to train an emulator that learned the relationship between the input parameter and the selection coefficient. We used a heteroskedastic Gaussian process (HGP) as an emulator, as this technique was highly efficient in previous analysis^5,16^. The HGP was trained using the function mleHetGP from the R-package hetGP^28^. Subsequently, we used the testing dataset to evaluate the accuracy of the emulator. To do that, we evaluated the correlation coefficient and root means squared error between the selection coefficients predicted by the emulator and the selection coefficients estimated by our model, as previously done in^5,16^. We then iteratively improved the emulator fit through adaptive sampling as in previous works^5,16^. Adaptive sampling involved sampling 100 parameter combinations from the parameter space region where we had lower confidence in the emulator’s predictions. This parameter space region was defined as the area where the emulator exhibited higher variance in its prediction. Once the 100 parameter combinations were sampled, the whole process was repeated (steps 2.1.1 to 2.1.3), leading to an improved fit of the emulator. After five rounds of adaptive sampling, each emulator achieved an excellent fit to the data (correlation coefficient above 0.98 for all emulators, see Supplementary Fig. 3 and 4).

##### 2.1.5 Global sensitivity analysis

After completing the final round of adaptive sampling, we used the emulator to assess the influence of each factor on the selection coefficient through global sensitivity analysis. The global sensitivity analysis was done using Sobol’s method of variance decomposition^21^. To do that, we first used Latin hypercube sampling to randomly sample two datasets of 100,000 parameter combinations. Then, we predicted the selection coefficient for each parameter combination using the emulators that could efficiently predict the selection coefficient for all these parameter combinations compared to our model. Finally, we performed the global sensitivity analysis on the datasets with 150,000 bootstrap replicates using the function soboljansen from the R-package Sensitivity^29^. This function returned the first-order indices of each factor, representing their relative influence on the selection coefficient (Fig. 2A and Supplementary Fig. 7 and 12), and assessed the 25th, 50th, and 75th quantiles of the predicted rate of spread for the two random datasets across each parameter range (Fig. 2B and Supplementary Fig. 9-11 and 13-20).

**Supplementary Table 8:** Parameters *and their ranges investigated in the global sensitivity analyses*.

| Determinant | Factors | Definitions | Ranges for vaccine aiming to prevent morbidity | Ranges for vaccine aiming to prevent transmission |
| --- | --- | --- | --- | --- |
| Vaccine Deployment | Coverage (%) | Percentage of individuals from the target age group that received SP-AQ during the first round of SMC (%) | 60-100 | 60-100 |
| Vaccine profile | Initial efficacy (%) | AIV: proportion of infections prevented from reaching the blood-stage (%).<br>BSV: proportion by which the multiplication rate of the parasite within-host is reduced (%).<br>TBV: proportion by which the probability of transmission from humans to mosquitoes is reduced (%). | 50 – 100 | 50 – 100 |
|  | Half-life (years) | Time for the initial efficacy to fall by 50% (years) | 0.5 – 1.5 | 0.5 – 5 |
| Biology | Degree of vaccine resistance (%) | Relative reduction of the initial efficacy of the resistant genotype compared to the sensitive one | 1-100 | 1-100 |
| Health systems | Access to treatment (%) | Percentage of symptomatic cases who received treatment within 14-days of symptom onset (%) | 10-80 | 10-80 |
| Transmission setting | EIR (inoculations per person per year) | The mean number of infective bites received by an individual during a year (inoculations per person per year) | 5-500 | 5-500 |

The choice of parameter range is described in section 2.1.1. SP-AQ=sulfadoxine-pyrimethamine+amodiaquine. AIV=anti-infective vaccine. BSV=blood-stage vaccine. TBV=transmission-blocking vaccine.

**Supplementary Figure 1:**
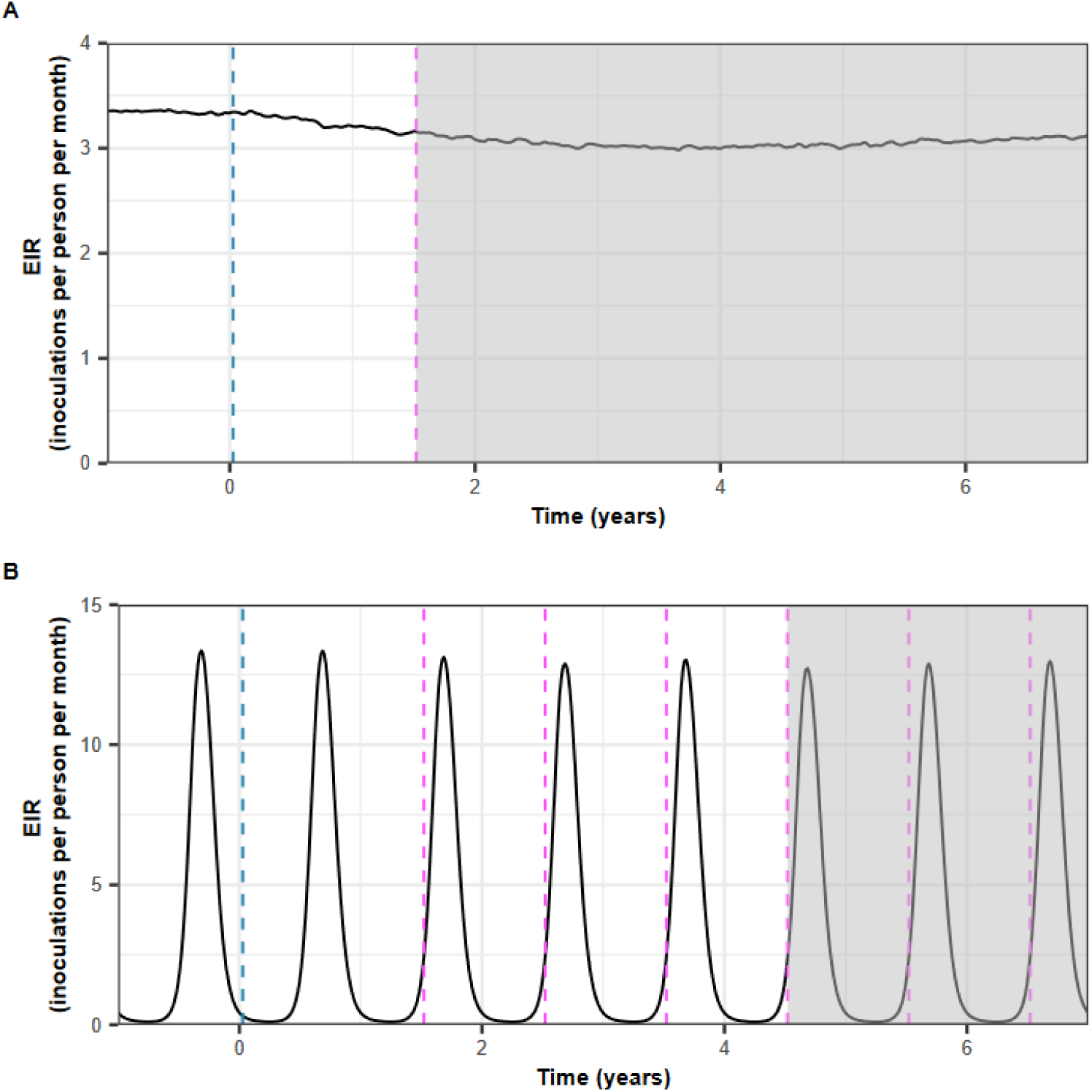
Illustration of the perennial and seasonal deployment strategies for vaccines that prevent morbidity in children. (**A**) The perennial deployment strategy for vaccines aiming to prevent morbidity in children. After the burn-in phase, the primary vaccination series was delivered to children at nine months of age, followed by a booster dose 18 months later. The blue line indicates when the first children received the primary vaccination series. The pink line indicates when children received the boosters. The grey area indicates the period when the rate of spread was estimated. (**B**) The seasonal deployment strategy of vaccines that aim to prevent morbidity in children. After the burn-in phase, a primary vaccination series was delivered to children at nine months of age. Children received a booster dose one month before the transmission pick in four consecutive years. To receive the booster before the transmission peak, children must have received the primary dose at least six months earlier. Year zero was defined as the first year during which the vaccine was deployed. The blue line indicates when the first children received the primary vaccination. The pink lines indicate when the children received the boosters. The grey area indicates the period when the spread rate was estimated. Both figures are visualised until year 7, but the spread was estimated until year 15 after the first deployment of the vaccine.

**Supplementary Figure 2:**
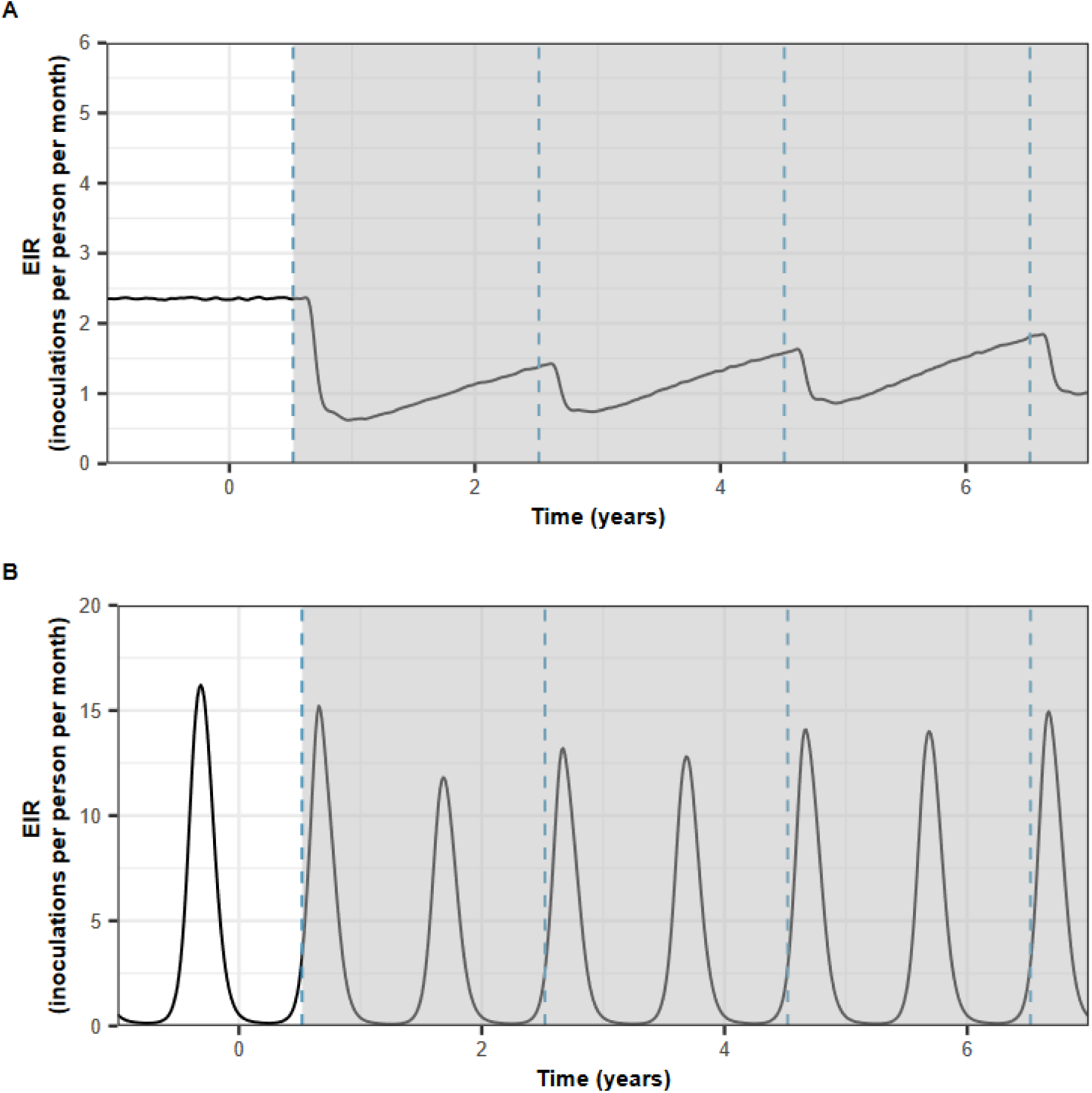
Illustration of the perennial and seasonal deployment strategies of vaccines that aim to reduce transmission. The strategies that aim to prevent transmission for (**A**) perennial and **(B**) seasonal vaccine deployment. After the burn-in phase, the vaccine was implemented in the entire population or administered to adults through mass administration, with a booster dose given every two years. In the seasonal setting, the mass administration occurred two months before the transmission peak. Year zero was defined as the first year during which the vaccine was deployed. The blue lines indicate when the rounds of vaccination were deployed. The grey areas indicate when the rate of spread could be estimated. Both figures stop at year 7, but the spread was estimated until year 15 after the first deployment of the vaccine.

**Supplementary Figure 3:**
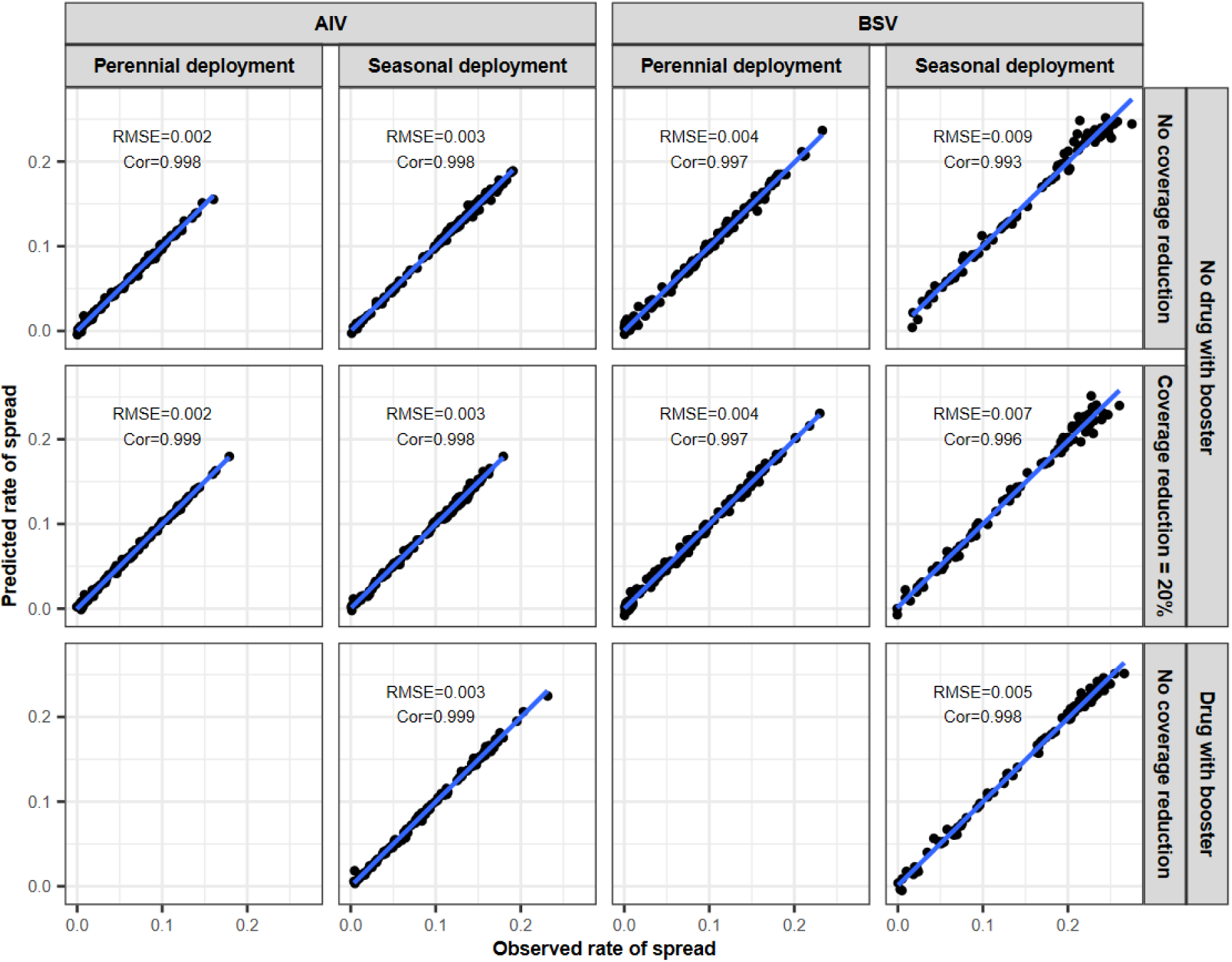
Accuracy of the emulators used for each global sensitivity analysis of the spread of genotypes resistant to an AIV or a BSV used to prevent morbidity in children. The figure displays the observed rate of spread estimated by our model simulations and the rate of spread predicted with the emulators for the testing dataset at the final round of adaptive sampling for each global sensitivity analysis. We performed the global sensitivity analyses for two different vaccine types (AIV and BSV), two different deployment strategies (perennial deployment and seasonal deployment), and different assumptions about the reduction of the coverage at the booster doses (0% or 20% from the initial vaccination). In the seasonal deployment, we either deployed the vaccine booster doses alone or with a drug combination. Combining vaccines with a drug was not done in the perennial setting as not currently considered. Cor=Spearman correlation coefficient, RMSE=root means squared error, blue lines: linear regression fit.

**Supplementary Figure 4:**
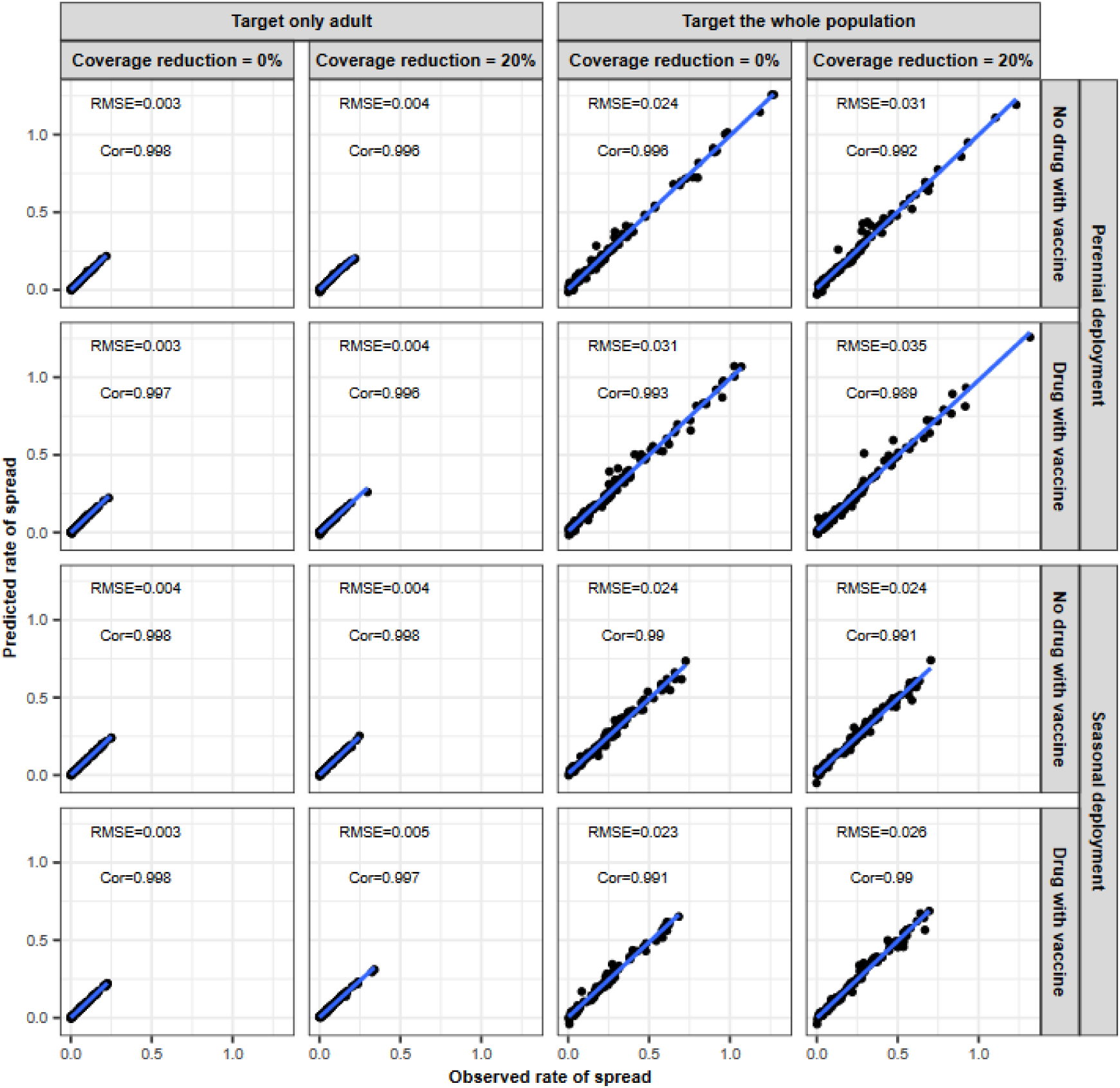
Accuracy of the emulators used for each global sensitivity analysis of the spread of genotype resistant to an AIV used to reduce transmission. The figure displays the observed rate of spread estimated by our model simulations and the rate of spread predicted with the emulators for the test dataset at the final round of adaptive sampling for each global sensitivity analysis. We performed the global sensitivity analyses when targeting two different age groups (adult or whole population) and deployed the vaccine with or without a drug in both perennial and seasonal settings. We assumed that the coverage remained constant or reduced by 20% from the previous round during the booster doses compared to the initial dose. Cor=Spearman correlation coefficient, RMSE=root means squared error, blue lines: linear regression fit.

#### 2.2 Conversion from the selection coefficient into the *T_50_*

To illustrate the impact of vaccine type and deployment strategy in a timeframe, we used the trained emulators to predict the selection coefficient for a set of parameter combinations and translated the selection coefficients to the time needed for the resistant genotype to spread from 1% to 50% frequency of inoculations, *T_50_* (Supplementary Note 2.2). Note that, in the real world, the resistant genotype could exist at a higher frequency than 1% leading to a shorter time to reach 50% frequency.

For each deployment strategy and vaccine type, we used the trained emulators to predict the selection coefficient of a genotype that has different degrees of resistance (0, 0.25, 0.5, 0.75, 1) to vaccines with different half-lives (1 or 1.5 years) and an initial efficacy of 95%. The vaccine was deployed at 90% coverage in a transmission setting with varying intensities of transmission (5 or 50 inoculations per person) and different levels of access to treatment (25% or 60%). Subsequently, we converted the selection coefficients into the numbers of parasite generations, *t*, needed for the frequency of the resistant genotype in inoculations to increase from 1% to 50% as follows^5,27^,

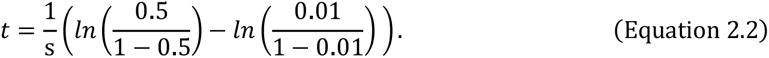

Then, the number of parasite generations was translated into the *T_50_*, assuming a parasite generation of 60 days^5,27^ (Fig. 3A and Supplementary Fig. 22 and 23). Supplementary Fig. 26B also display the results when we calculated the time needed for the resistant genotype to spread from 1% to 50% frequency of inoculations, *T_100_*.

**Supplementary Figure 5:**
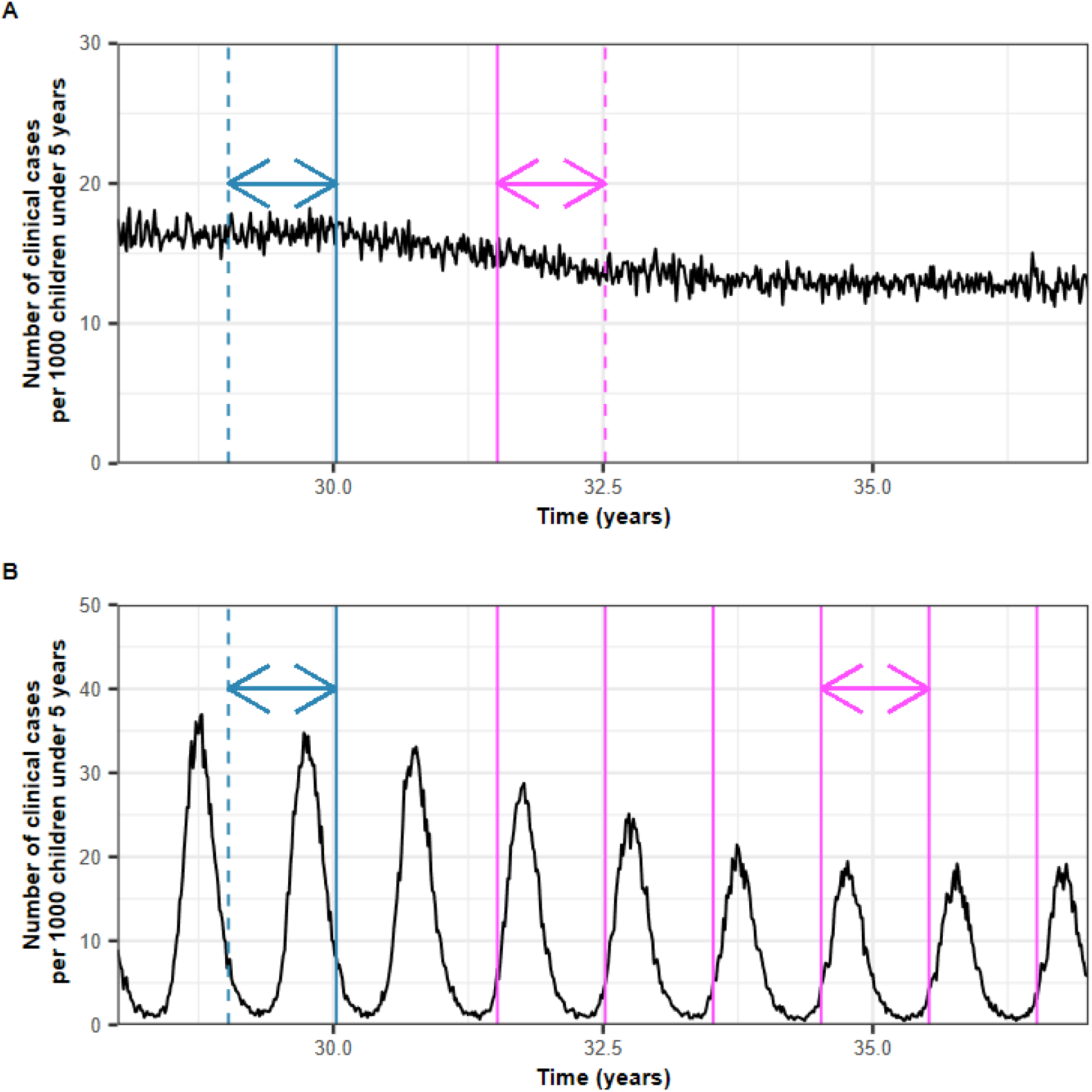
An illustration of the estimate for the effectiveness of vaccines that aim to reduce morbidity in children. (**A**) The figures illustrate the estimation of the effectiveness of vaccines that aim to reduce morbidity in the perennial setting. The black curve represents the incidence of malaria clinical cases per 1,000 children under five years old. The solid blue line represents the first time the primary vaccination series was deployed to a child. The blue arrow indicates the period during which we estimated the incidence rate of clinical malaria before the vaccine deployment. The solid pink line represents the first time a booster dose was given to a child. The pink double-ended arrow represents the period during which we estimated the incidence of clinical malaria during vaccine implementation. (**B**) The figures illustrate the estimate for the effectiveness of vaccines that aim to reduce morbidity in the seasonal setting. The black curve represents the incidence of malaria clinical cases per 1,000 children under five years old. The solid blue line represents the first time the primary vaccination series was deployed to a child. The blue double-ended arrow indicates the period where we estimated the incidence rate of clinical malaria before vaccine deployment. The solid pink lines represent the yearly deployment of vaccine boosters. The pink double-ended arrow represents the period during which we estimated the incidence of clinical malaria during vaccine implementation.

**Supplementary Figure 6:**
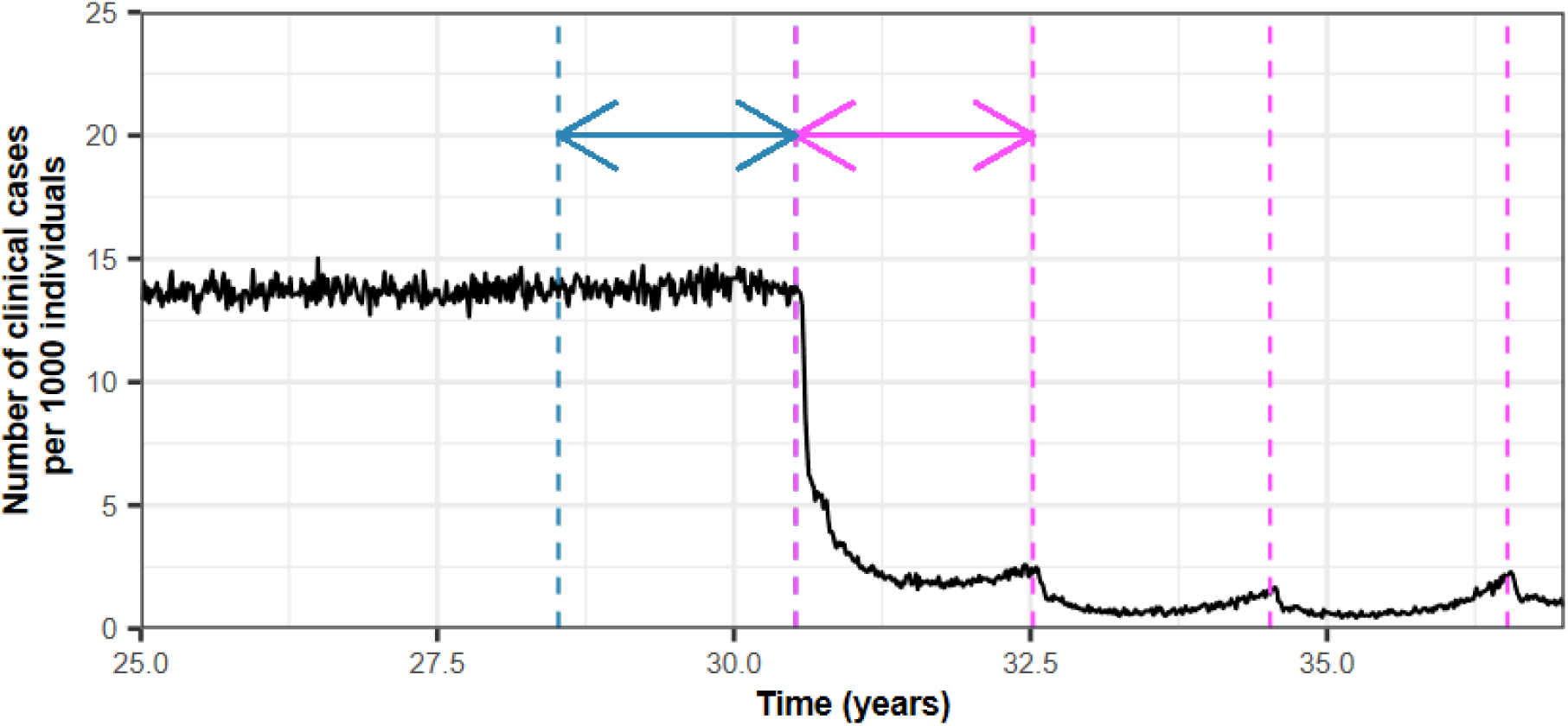
An illustration of the estimate for the effectiveness of vaccines that aim to reduce the transmission. The black curves represent the incidence of malaria clinical cases per 1,000 individuals. The pink dashed lines represent the deployment of the vaccine to the whole population. The pink double-ended arrow represents the period during which we estimated the incidence of clinical malaria during the first two years of vaccine implementations. The blue double-ended arrow and dashed lines indicate the period during which we estimated the incidence rate of clinical malaria before the deployment of the vaccine.

**Supplementary Figure 7:**
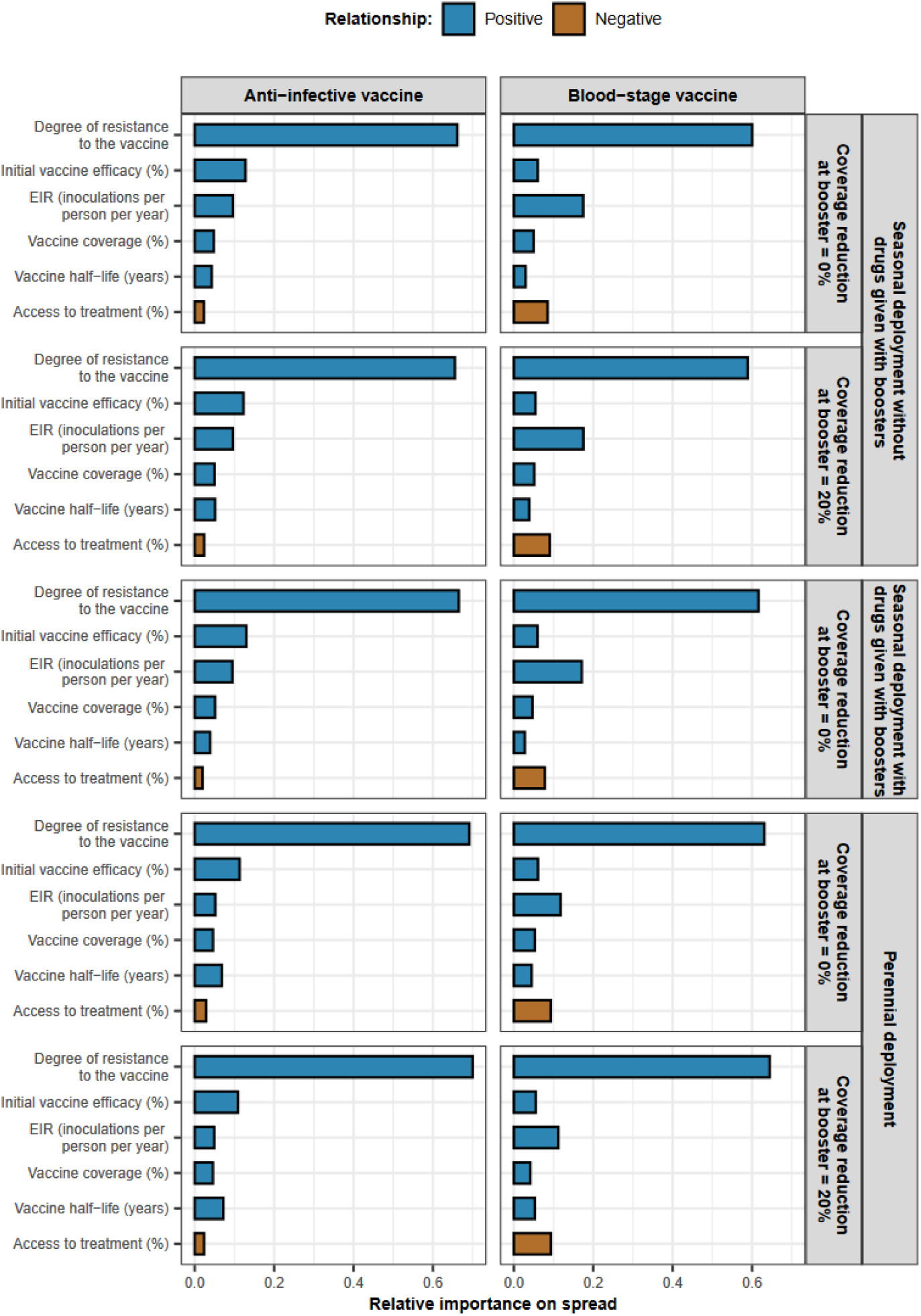
The relative contribution of each factor on the spread of vaccine-resistant genotypes for each deployment strategy and each vaccine type that aimed to prevent morbidity in children. The relative contribution of each factor was assessed through global sensitivity analysis when an AIV or a BSV was deployed to children in a perennial or seasonal setting. We assumed that the coverage of the vaccine was constant or decreased by 20% between the primary vaccination and each booster. For the seasonal deployment, we deployed the booster dose alone or with a drug. As deploying vaccines with drugs via EPI is not recommended, we did not explore dual administration in perennial settings. Factors in blue and brown represent a positive and negative relationship with the rate of spread, respectively. The parameter ranges and definitions are described in Supplementary Table 8.

**Supplementary Figure 8:**
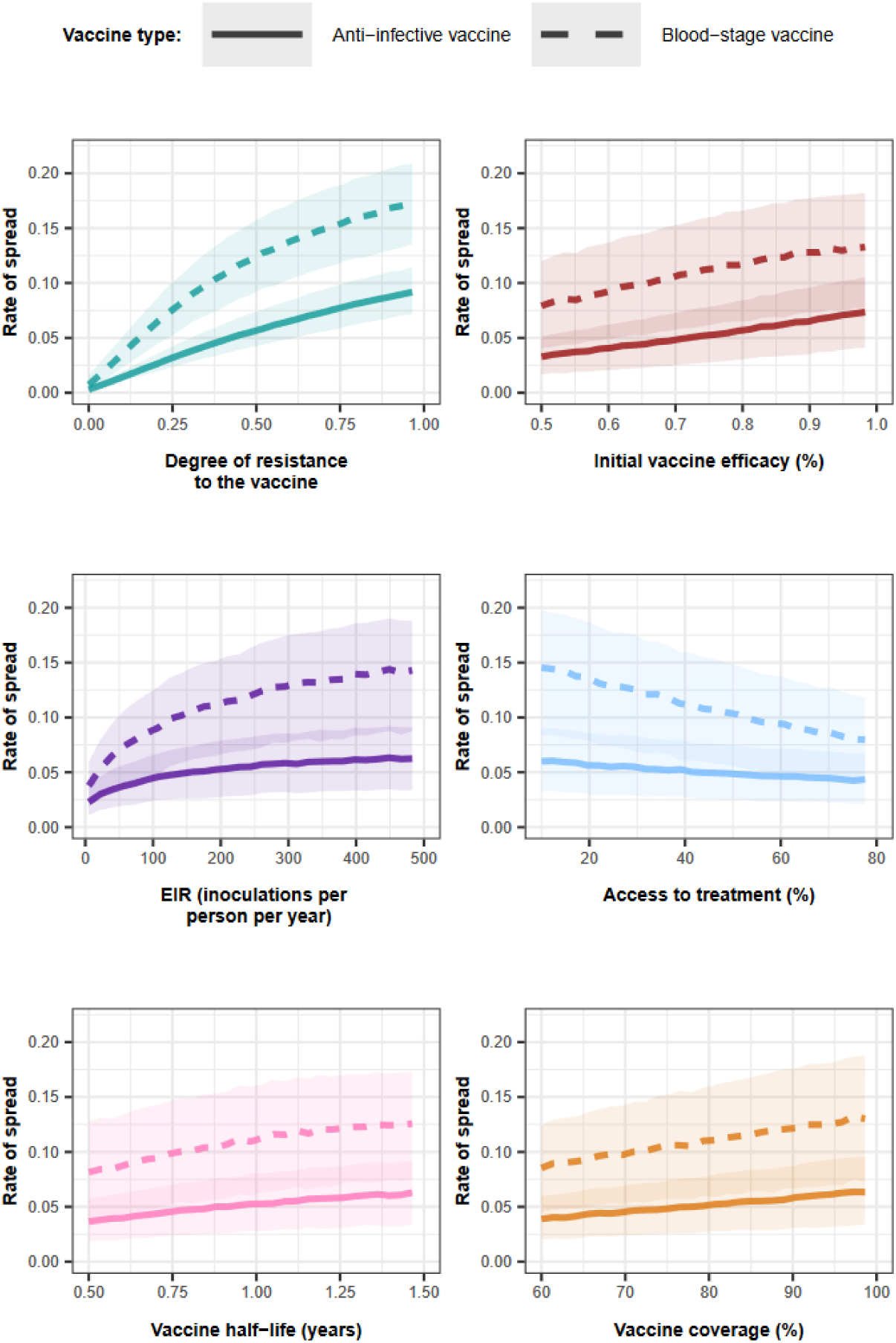
Influence of all factors on the rate of spread of a genotype resistant to an AIV or a BSV that aimed to prevent morbidity when vaccines were implemented in the seasonal setting with coverage that decreased by 20% at the booster dose and no drug was given with the booster doses. Curves and shaded areas represent the median and interquartile range of the rate of spread of a genotype resistant to an AIV (solid curve) or a BSV (dashed curve) estimated during the global sensitivity analyses across the parameter range described in Supplementary Table 8.

**Supplementary Figure 9:**
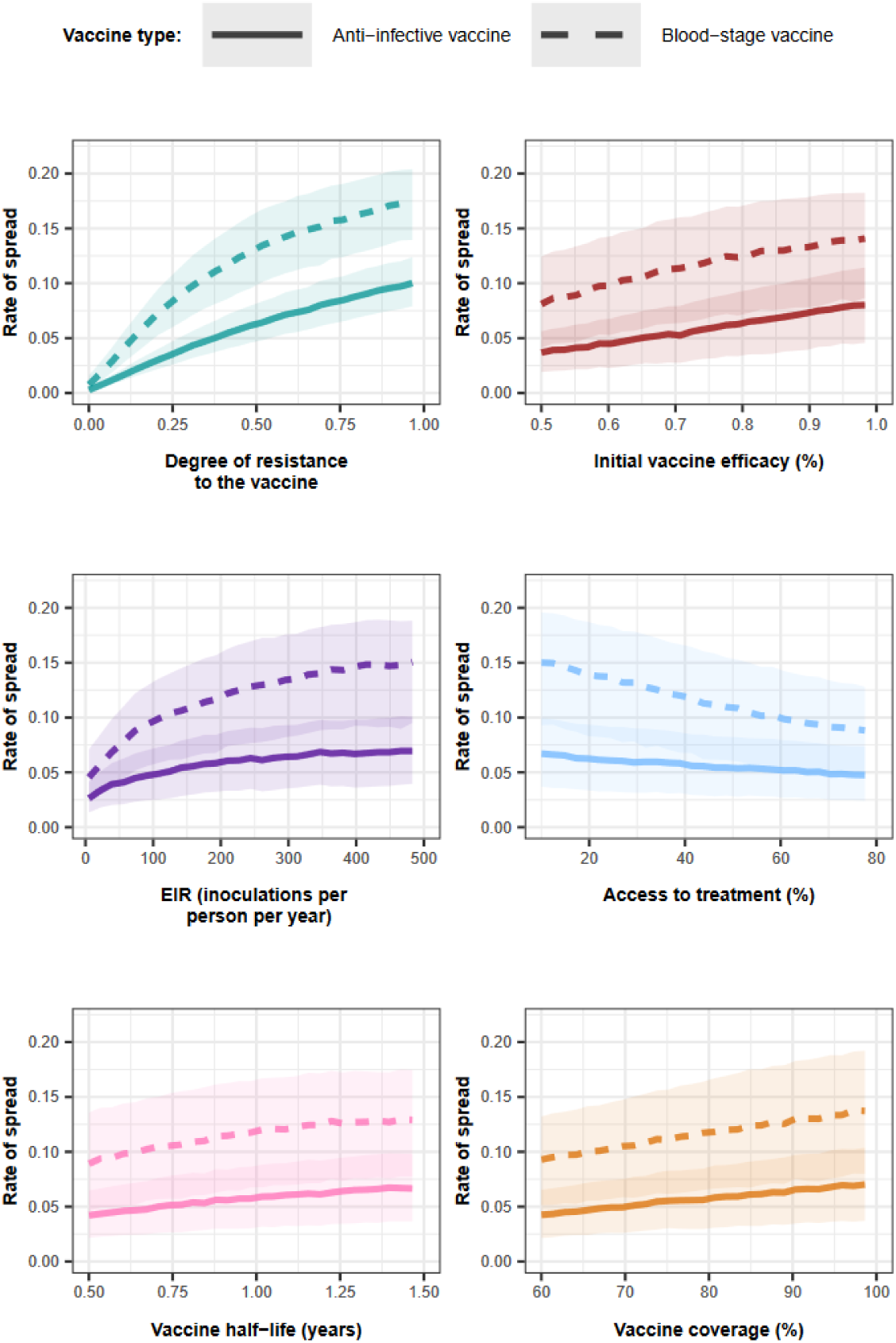
Influence of all factors on the rate of spread of a genotype resistant to an AIV or a BSV that aimed to prevent morbidity when vaccines were implemented in the seasonal setting with coverage that did not decrease at the booster dose and a drug was given with the booster doses. Curves and shaded areas represent the median and interquartile range of the rate of spread of a genotype resistant to an AIV (solid curve) or a BSV (dashed curve) estimated during the global sensitivity analyses across the parameter range described in Supplementary Table 8.

**Supplementary Figure 10:**
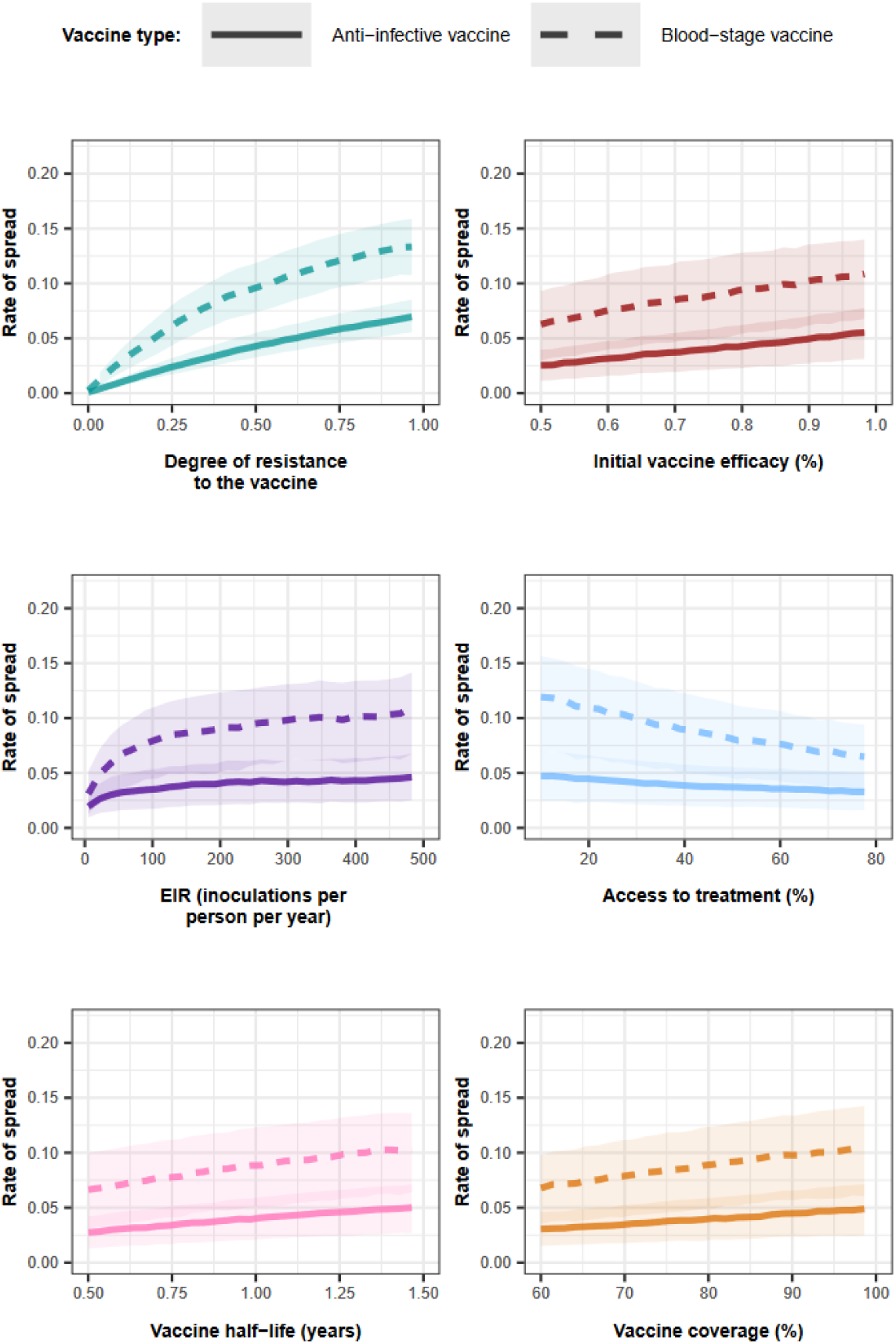
Influence of all factors on the rate of spread of a genotype resistant to an AIV or a BSV that aimed to prevent morbidity when vaccines were implemented in the perennial setting with coverage that did not decrease at the booster dose. Curves and shaded areas represent the median and interquartile range of the rate of spread of a genotype resistant to an AIV (solid curve) or a BSV (dashed curve) estimated during the global sensitivity analyses across the parameter range described in Supplementary Table 8.

**Supplementary Figure 11:**
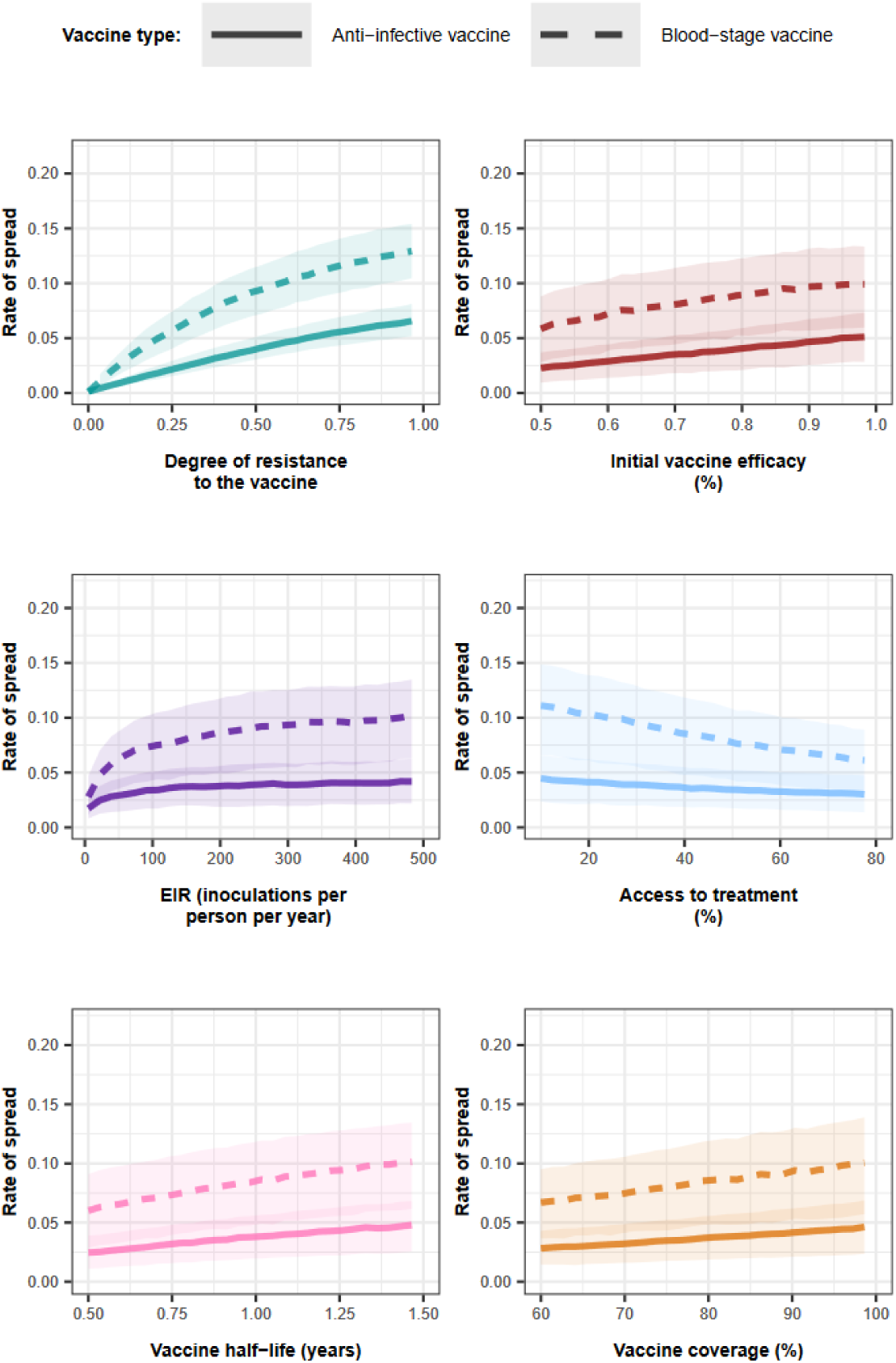
Influence of all factors on the rate of spread of a genotype resistant to an AIV or a BSV that aimed to prevent morbidity when vaccines were implemented in the perennial setting with coverage that decreased by 20% at the first booster. Curves and shaded areas represent the median and interquartile range of the rate of spread of a genotype resistant to an AIV (solid curve) or a BSV (dashed curve) estimated during the global sensitivity analyses across the parameter range described in Supplementary Table 8.

**Supplementary Figure 12:**
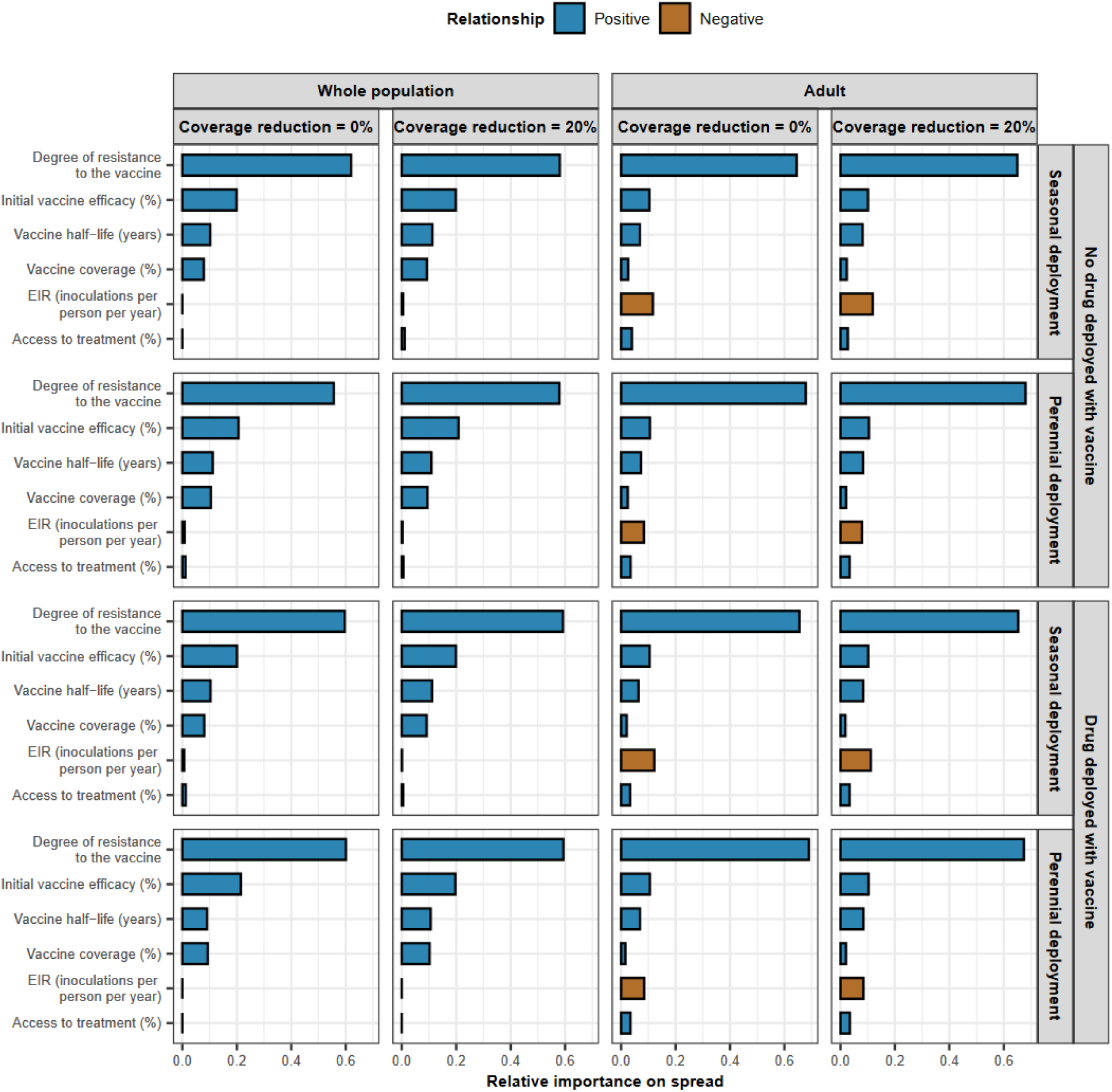
The relative contribution of each factor in determining the spread of vaccine-resistant genotype for each deployment strategy for vaccines that reduce transmission. The relative contribution of each factor was assessed through global sensitivity analysis when an AIV was deployed to the whole population or adults. The AIV was deployed with or without a drug in a perennial or seasonal setting. We assumed that the vaccine coverage was constant or decreased by 20% between the primary vaccination and each booster dose. Factors in blue and brown represent a positive and negative relationship with the rate of spread, respectively. The parameter ranges and definitions are described in Supplementary Table 8.

**Supplementary Figure 13:**
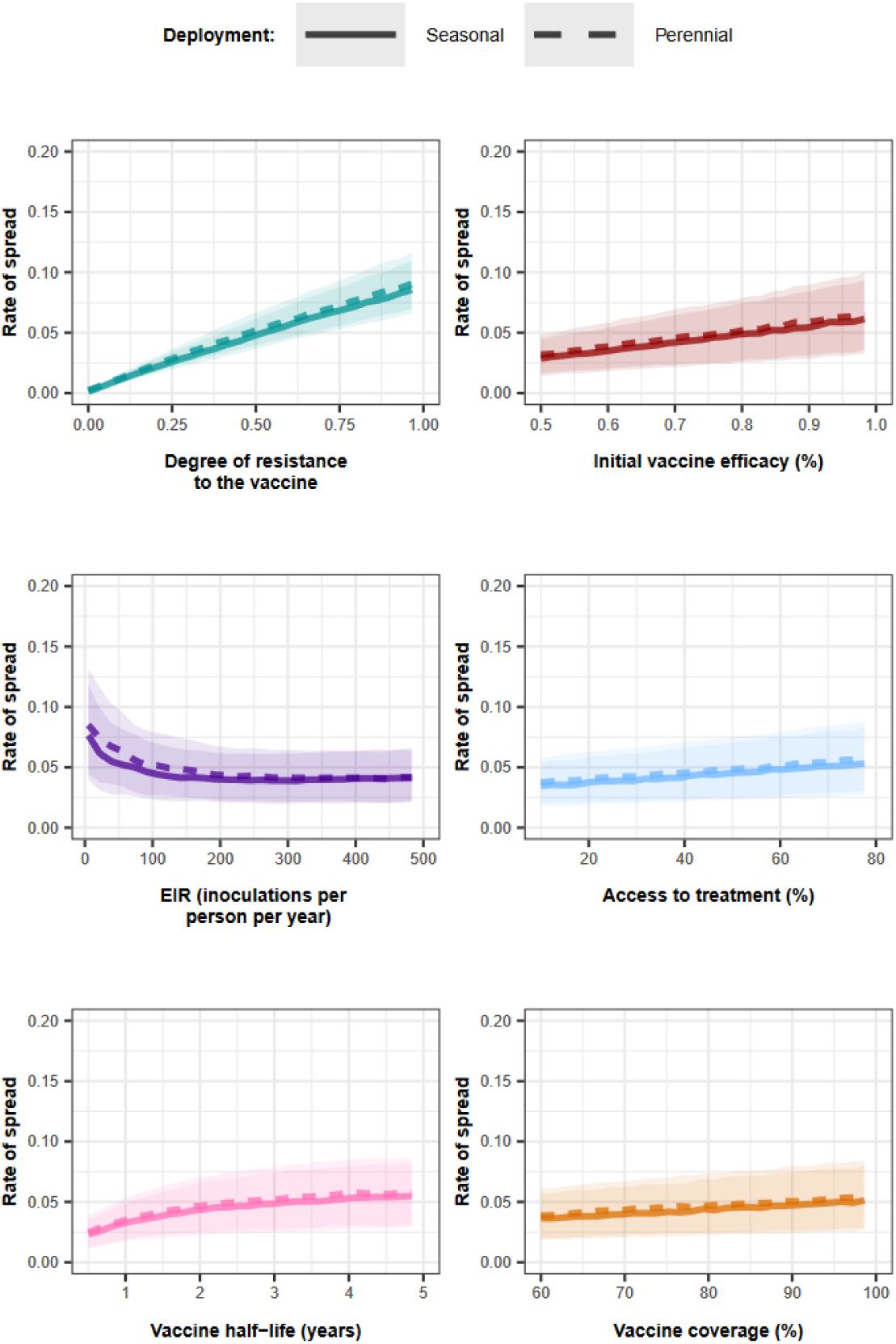
Influence of all factors on the rate of spread of genotype resistant to an AIV that aimed to reduce transmission when the vaccine was deployed to adults without a drug, and the coverage decreased by 0% at the booster dose. Curves and shaded areas represent the median and interquartile range of the rate of spread of a genotype resistant to an AIV in a seasonal (solid curve) or a perennial (dashed curve) transmission settings estimated during the global sensitivity analyses across the parameter ranges described in Supplementary Table 8.

**Supplementary Figure 14:**
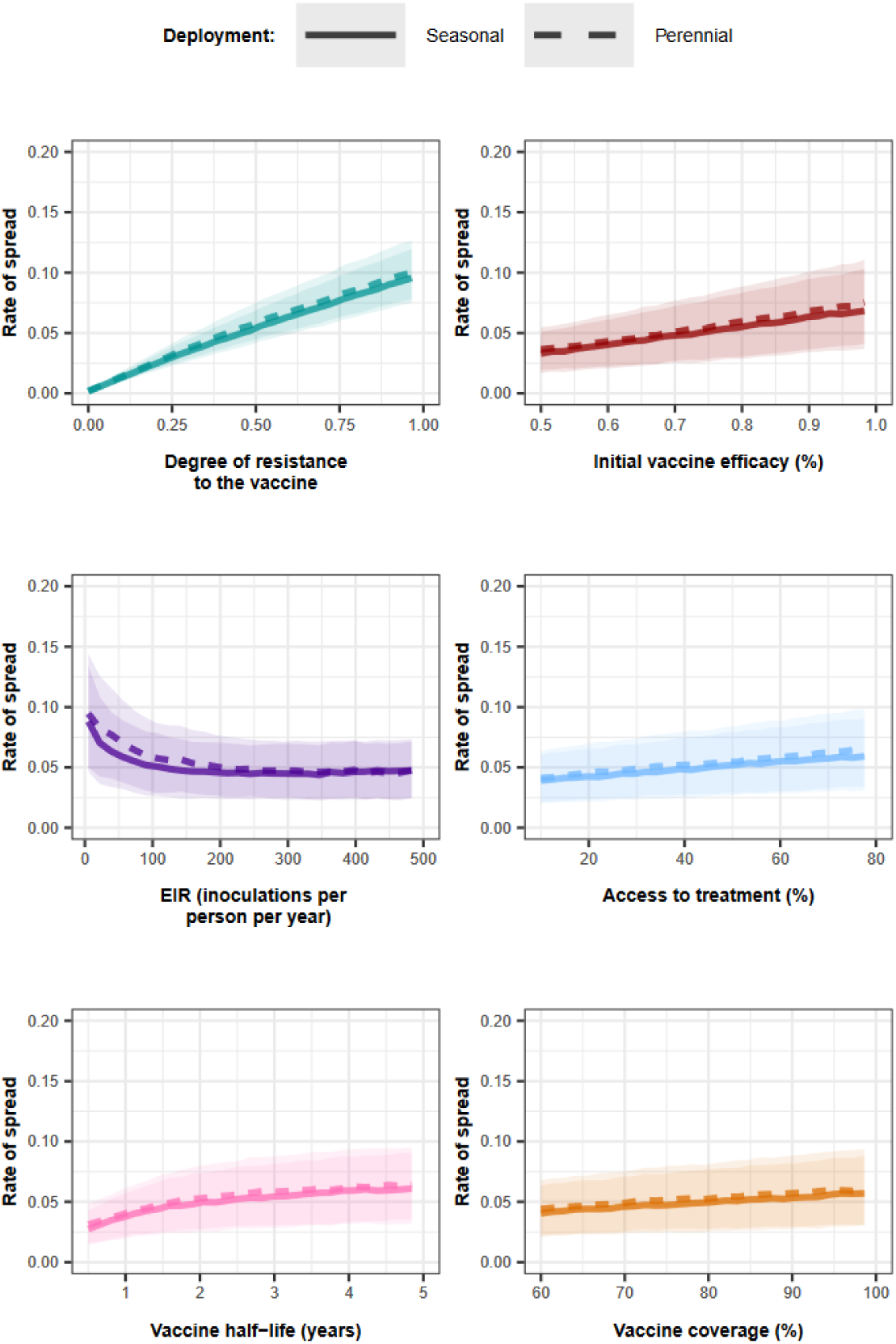
Influence of all factors on the rate of spread of genotype resistant to an AIV that aimed to reduce transmission when the vaccine was deployed to adults without a drug, and the coverage decreased by 20% at the booster dose. Curves and shaded areas represent the median and interquartile range of the rate of spread of a genotype resistant to an AIV in a seasonal (solid curve) or a perennial (dashed curve) transmission settings estimated during the global sensitivity analyses across the parameter ranges described in Supplementary Table 8.

**Supplementary Figure 15:**
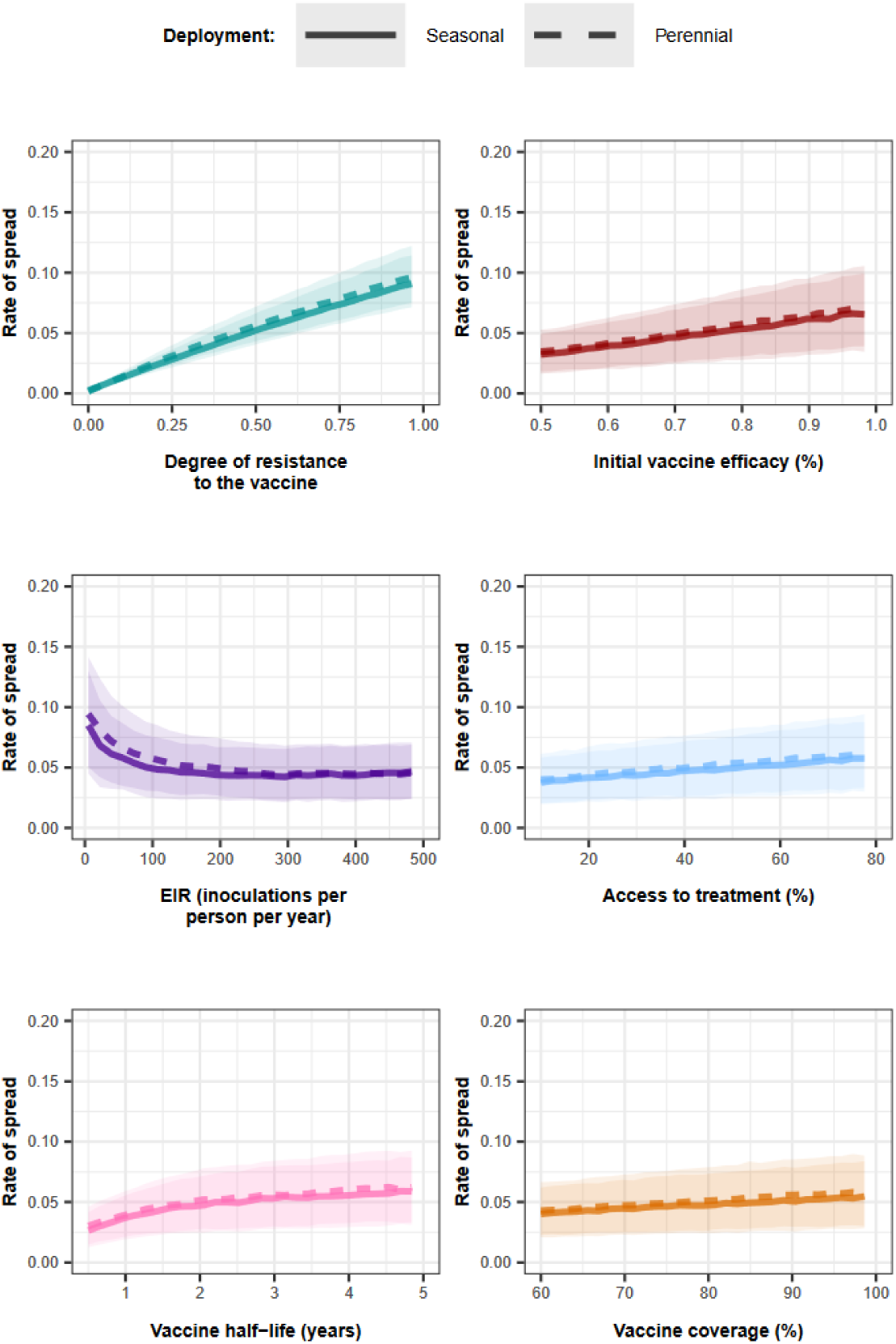
Influence of all factors on the rate of spread of a genotype resistant to an AIV that aimed to reduce transmission when the vaccine was deployed to adults with a drug, and the coverage did not decrease at the booster dose. Curves and shaded areas represent the median and interquartile range of the rate of spread of a genotype resistant to an AIV in a seasonal (solid curve) or a perennial (dashed curve) transmission settings estimated during the global sensitivity analyses across the parameter ranges described in Supplementary Table 8.

**Supplementary Figure 16:**
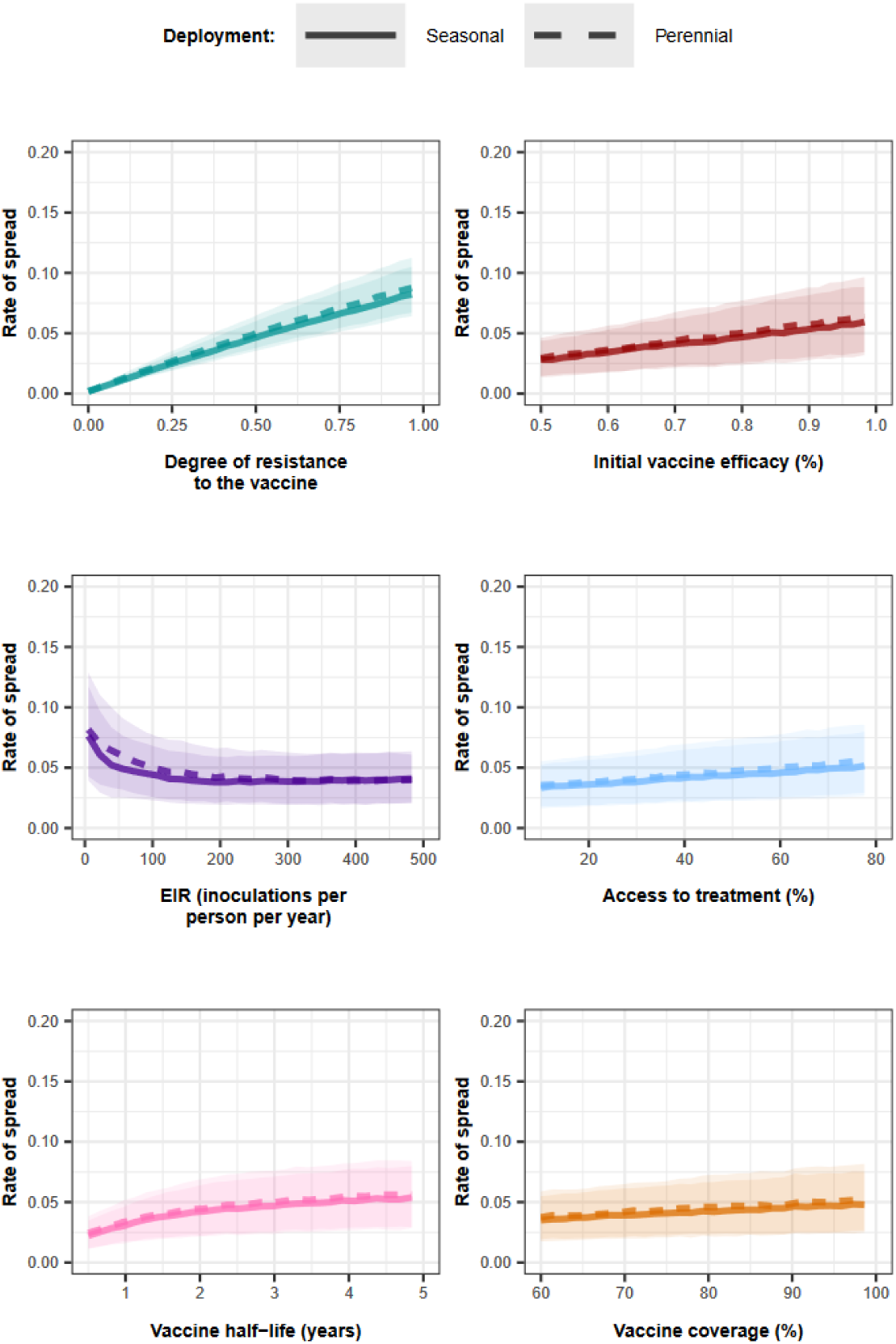
Influence of all factors on the rate of spread of a genotype resistant to an AIV that aimed to reduce transmission when the vaccine was deployed to adults with a drug, and the coverage decreased by 20% at the booster dose. Curves and shaded areas represent the median and interquartile range of the rate of spread of a genotype resistant to an AIV in a seasonal (solid curve) or a perennial (dashed curve) transmission settings estimated during the global sensitivity analyses across the parameter ranges described in Supplementary Table 8.

**Supplementary Figure 17:**
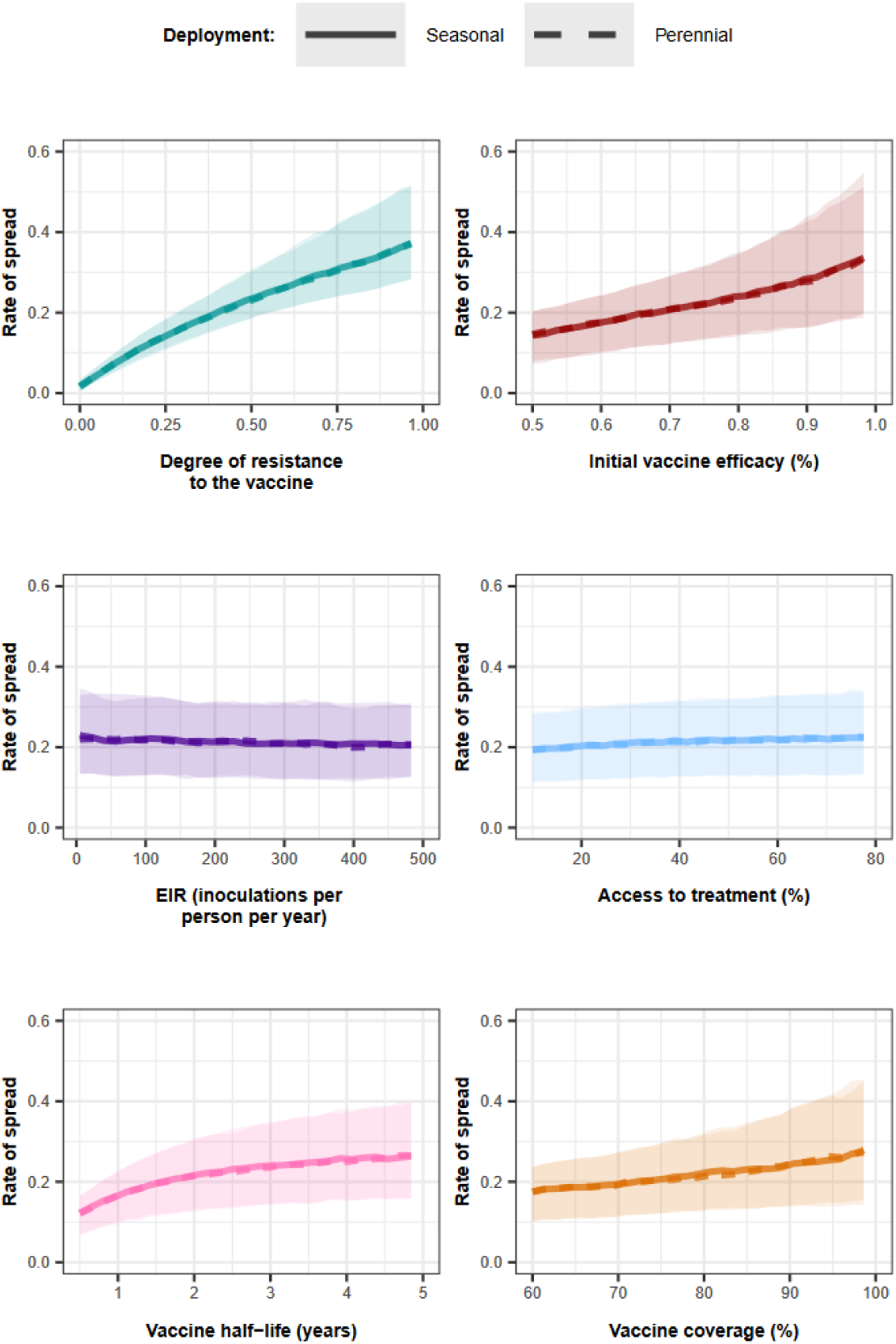
Influence of all factors on the rate of spread of genotype resistant to an AIV that aimed to reduce transmission when the vaccine was deployed to the whole population without a drug, and the coverage did not decrease at the booster dose. Curves and shaded areas represent the median and interquartile range of the rate of spread of a genotype resistant to an AIV in a seasonal (solid curve) or a perennial (dashed curve) transmission settings estimated during the global sensitivity analyses across the parameter ranges described in Supplementary Table 8.

**Supplementary Figure 18:**
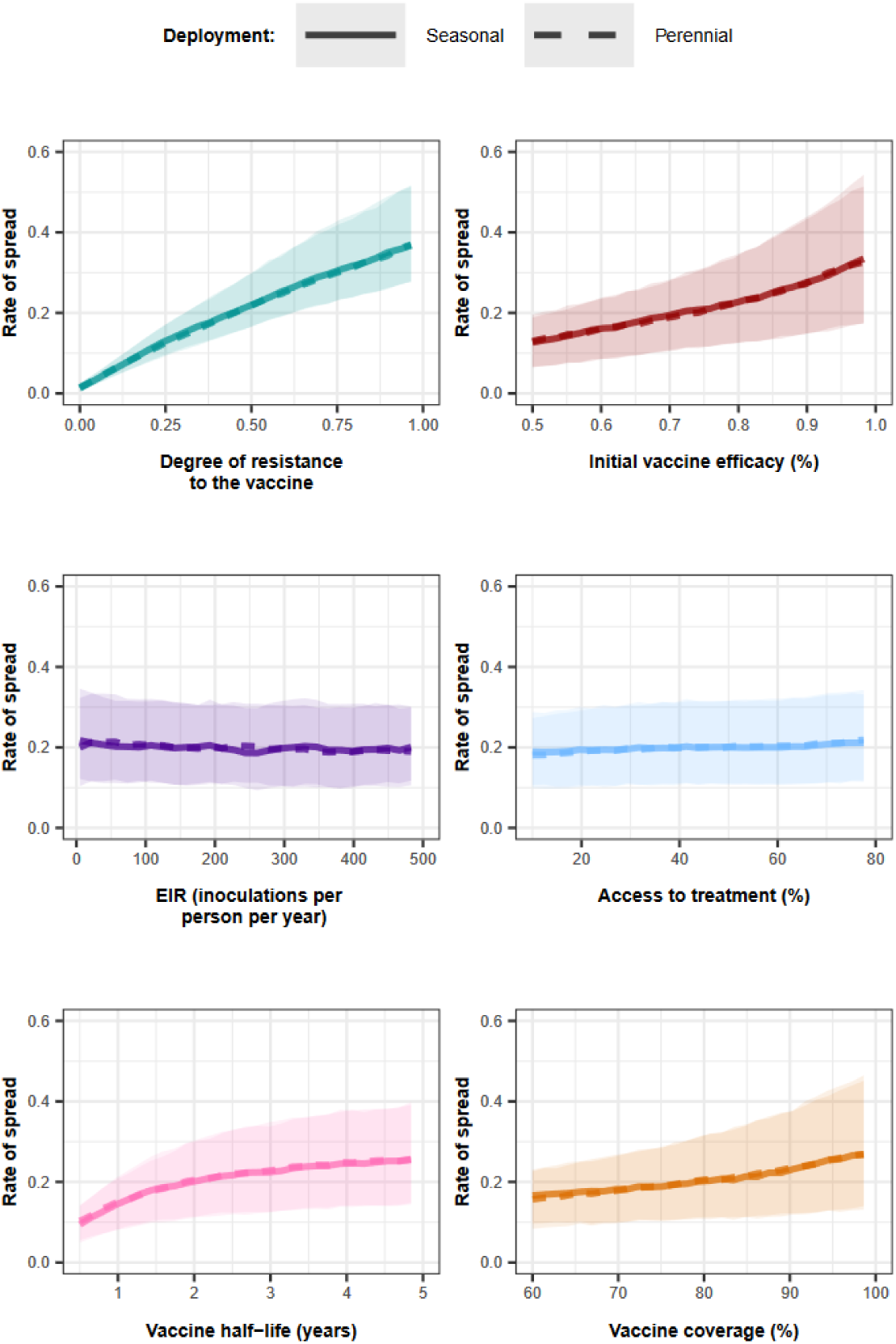
Influence of all factors on the rate of spread of genotype resistant to an AIV that aimed to reduce transmission when the vaccine was deployed to the whole population without a drug, and the coverage decreased by 20% at the booster dose. Curves and shaded areas represent the median and interquartile range of the rate of spread of a genotype resistant to an AIV in a seasonal (solid curve) or a perennial (dashed curve) transmission settings estimated during the global sensitivity analyses across the parameter ranges described in Supplementary Table 8.

**Supplementary Figure 19:**
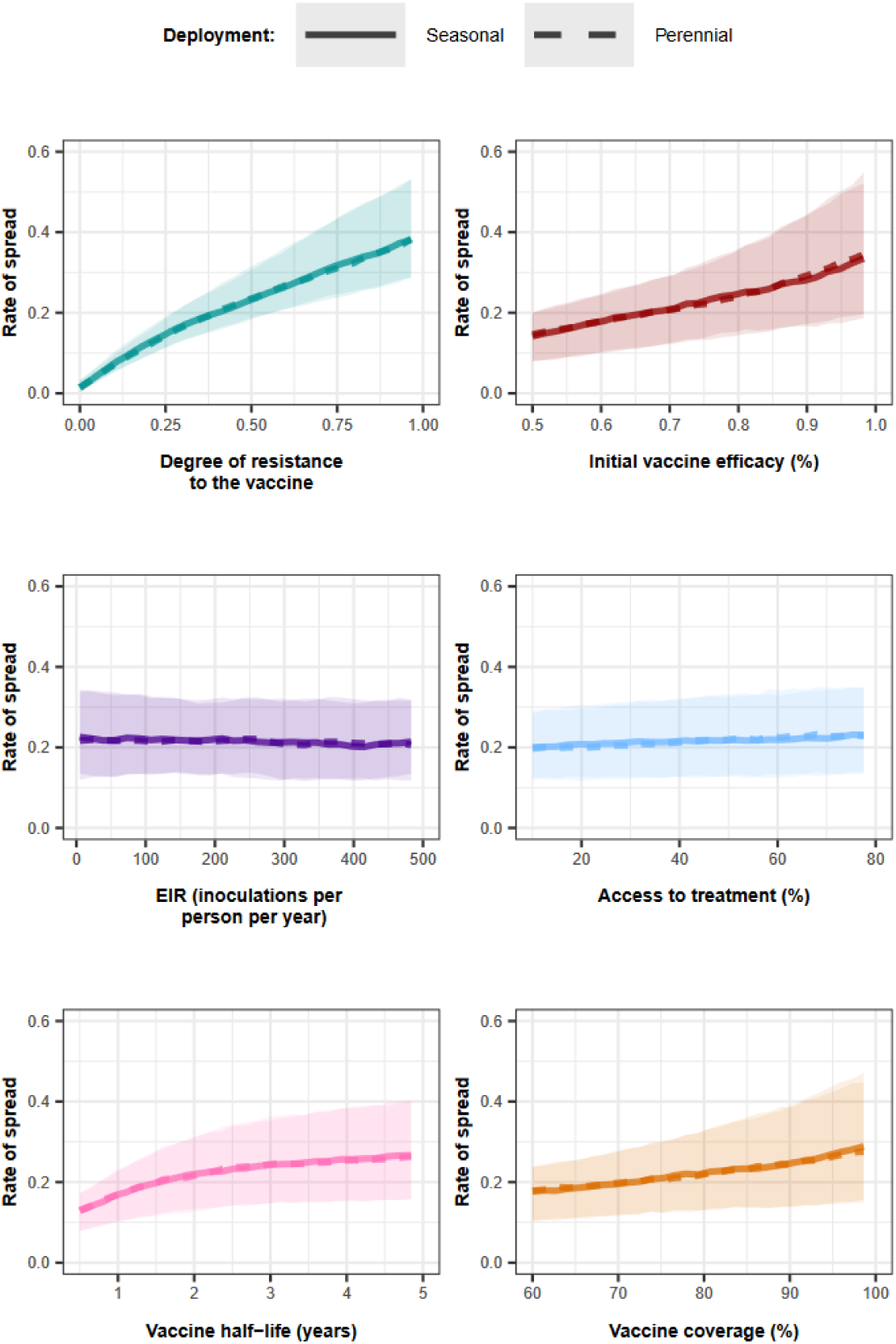
Influence of all factors on the rate of spread of genotype resistant to an AIV that aimed to reduce transmission when the vaccine was deployed to the whole population with a drug, and the coverage did not decrease at the booster dose. Curves and shaded areas represent the median and interquartile range of the rate of spread of a genotype resistant to an AIV in a seasonal (solid curve) or a perennial (dashed curve) transmission settings estimated during the global sensitivity analyses across the parameter range described in Supplementary Table 8.

**Supplementary Figure 20:**
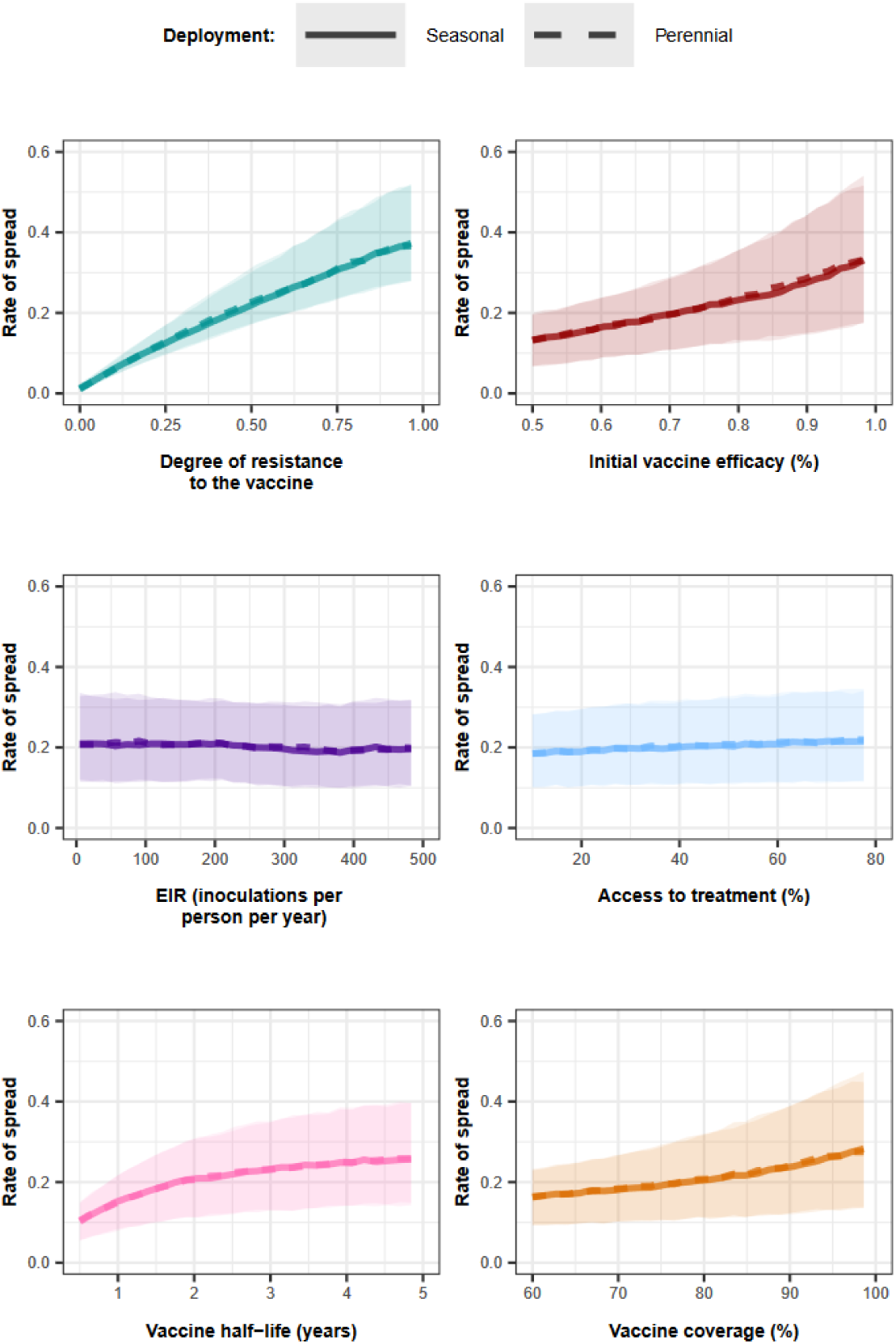
Influence of all factors on the rate of spread of genotype resistant to an AIV that aimed to reduce transmission when the vaccine was deployed to the whole population with a drug, and the coverage decreased by 20% at the booster dose. Curves and shaded areas represent the median and interquartile range of the rate of spread of a genotype resistant to an AIV in a seasonal (solid curve) or a perennial (dashed curve) transmission settings estimated during the global sensitivity analyses across the parameter ranges described in Supplementary Table 8.

**Supplementary Figure 21:**
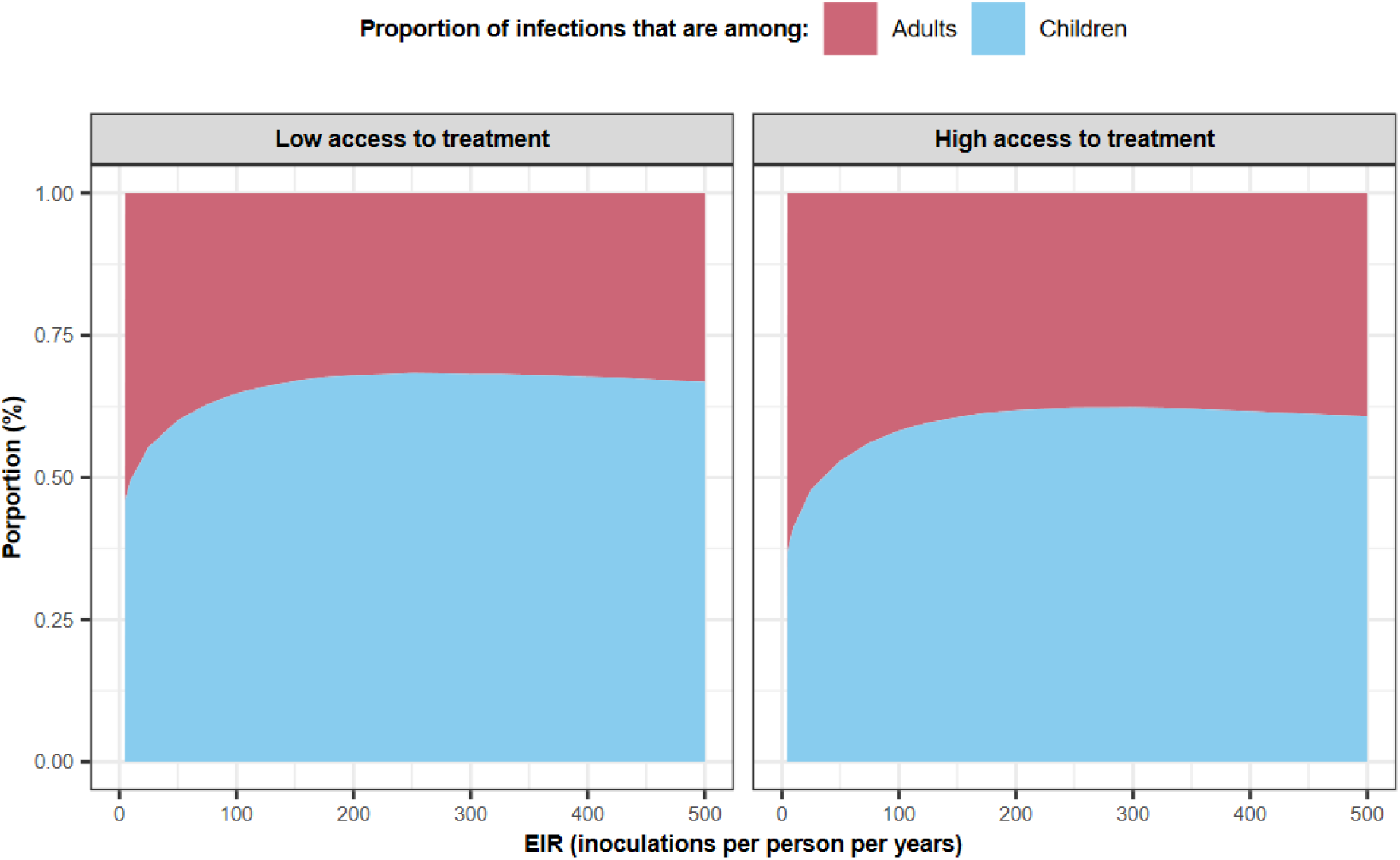
Proportion of infections among adults and children. The relationship between the EIR (inoculations per person per year) and the proportion of infections detectable by microscopy that are among adults above 18 years old (red area) and children under 18 years old (blue area) for different degrees of access to treatment (low access to treatment=25%, high access to treatment=70%).

**Supplementary Figure 22:**
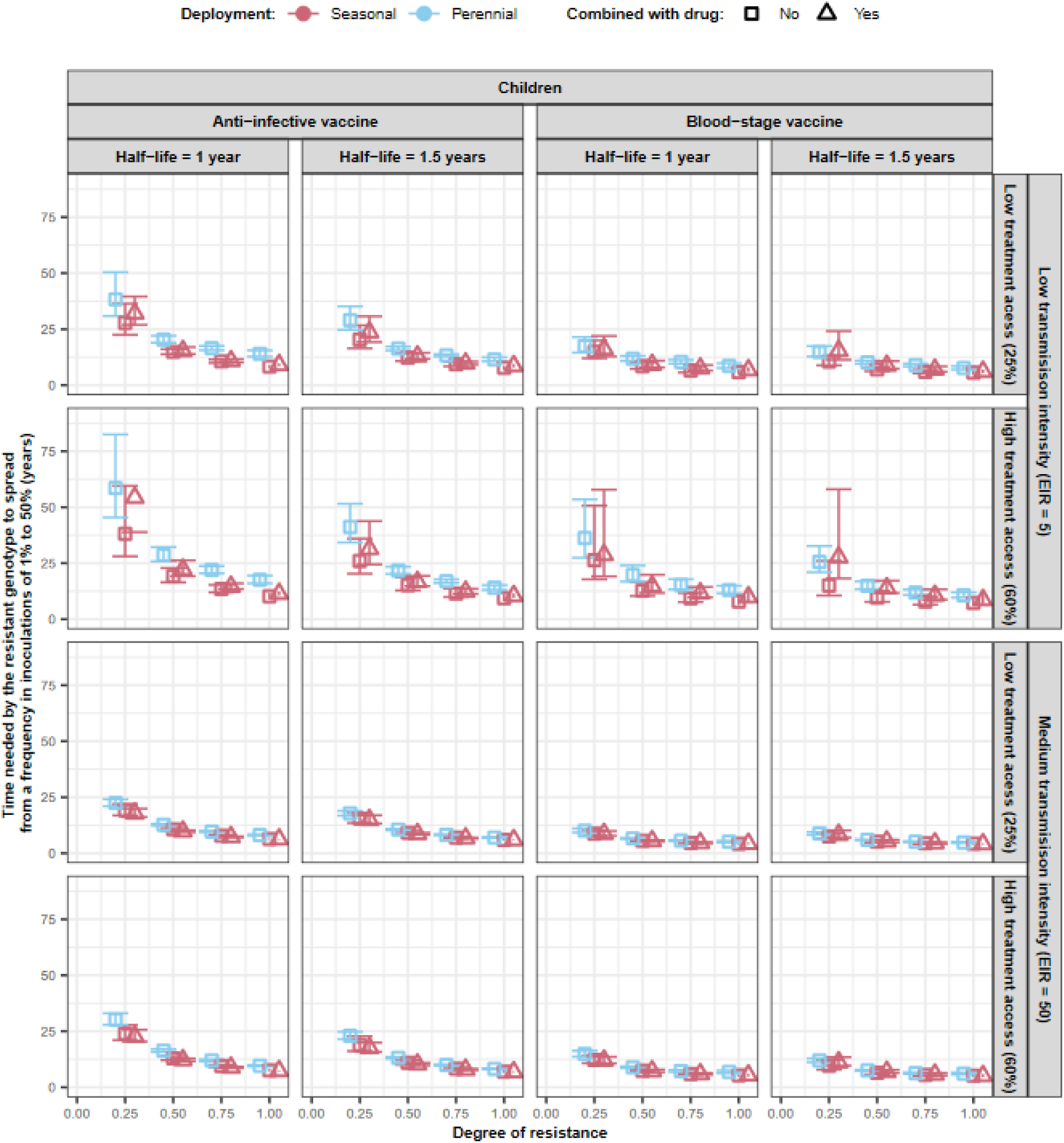
The impact of the degree of resistance on the number of years until the resistant genotype is dominant for vaccines that reduce morbidity in children The predicted time needed for genotypes with different degrees of resistance (0, 0.25, 0.5, 0.75, 1) to vaccines to spread from a frequency in inoculations of 1% to 50%, *T_50_*. We predicted the *T_50_* and 95% confidence intervals for each vaccine type and deployment strategy **that aims to reduce morbidity in children** (perennial deployment (light red marker), seasonal deployment (light blue marker), with a drug (triangle), or without a drug (square) implemented with the vaccines. We evaluated the *T_50_* for vaccines with an initial efficacy of 95% and a half-life of 1.0 or 1.5 years. These vaccines were deployed with a coverage of 90% that did not decrease at the booster doses in settings with an EIR of 5 or 50 inoculations per person per year and low (25%) or high (60%) levels of access to treatment.

**Supplementary Figure 23:**
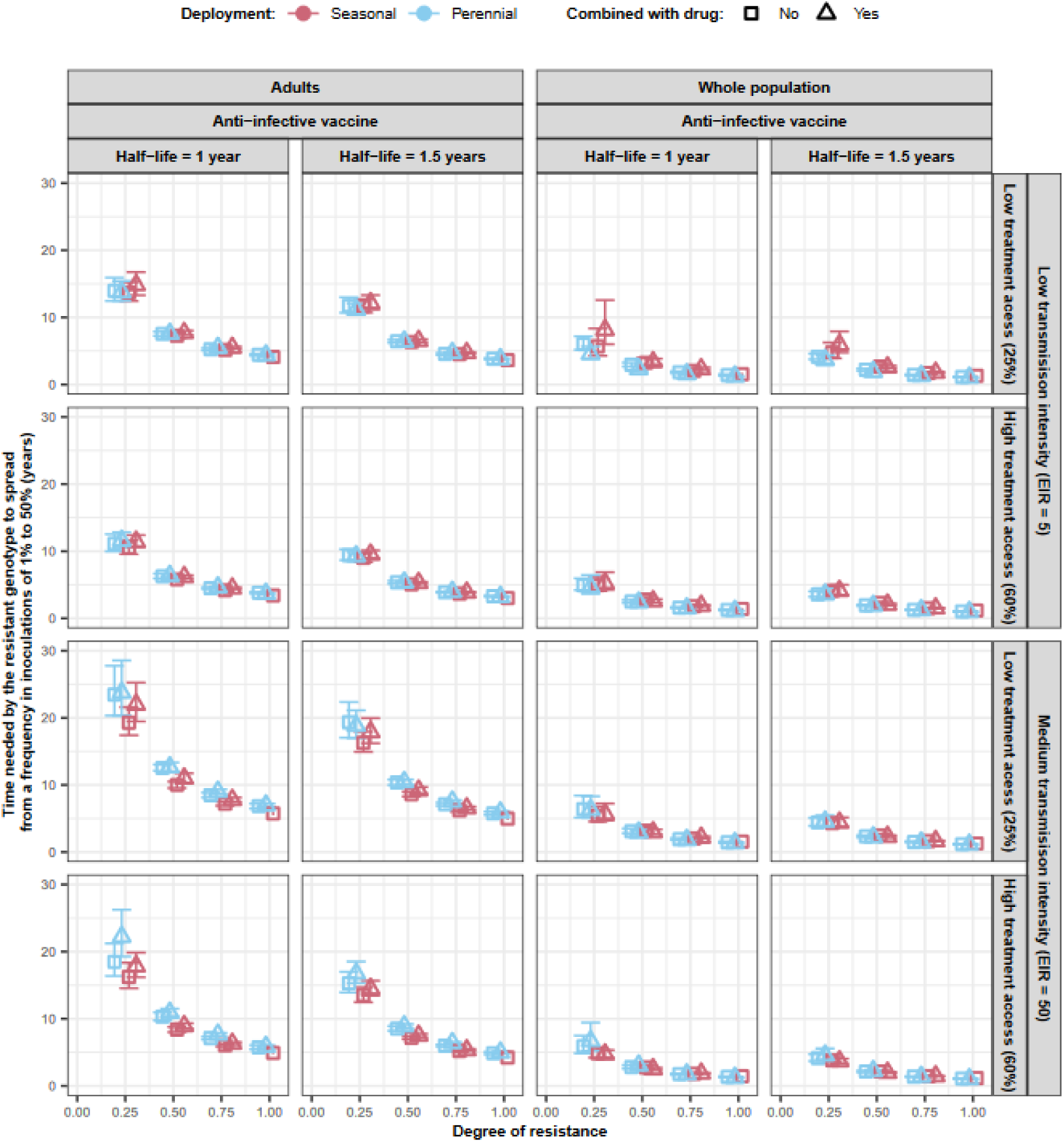
The impact of the degree of resistance on time needed before the resistant genotype becomes dominant for vaccines that reduce transmission The predicted time needed for genotypes with different degrees of resistance (0, 0.25, 0.5, 0.75, 1) to vaccines to spread from a frequency in inoculations of 1% to 50%, *T_50_*. We predicted the *T_50_* and 95% confidence intervals for each vaccine type and deployment strategy **that aims to reduce transmission** (perennial deployment (light red marker), seasonal deployment (light blue marker), with a drug (triangle), or without a drug (square) implemented with the vaccines. We evaluated the *T_50_* for vaccines with an initial efficacy of 95% and a half-life of 1.0 or 1.5 years. These vaccines were deployed with a coverage of 90% that did not decrease at the booster doses in settings with an EIR of 5 or 50 inoculations per person per year and low (25%) or high (60%) levels of access to treatment.

**Supplementary Figure 24:**
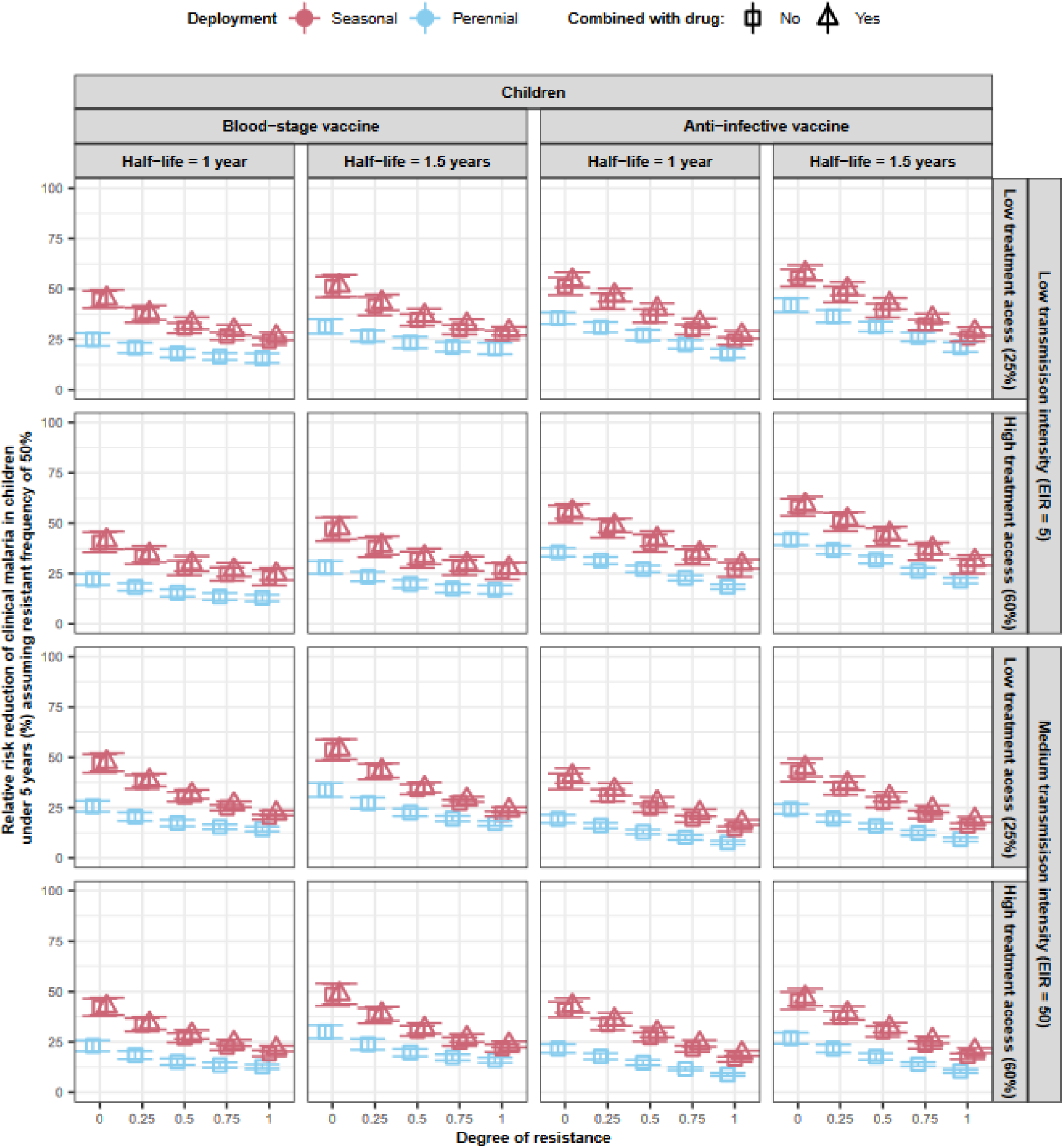
The impact of the degree of resistance on the effectiveness of vaccines that reduce morbidity in children. Estimated vaccine effectiveness (VE) of the vaccine in a population composed of 50% sensitive genotypes and 50% resistant genotypes with different degrees of resistance (0, 0.25, 0.5, 0.75, 1) to vaccines **that aimed to reduce morbidity in children**. The VE was estimated as the relative reduction in the number of clinical malaria cases among children under five years of age. The VE and 95% confidence intervals were predicted for each vaccine type and deployment strategy (perennial deployment (light red light marker), seasonal deployment (light blue marker), with a drug (triangle), or without a drug (square) implemented with the vaccines. We evaluated the VE for vaccines with an initial efficacy of 95% and a half-life of 1.0 or 1.5 years. These vaccines were deployed with a coverage of 90% that did not decrease at the booster doses in settings with an EIR of 5 or 50 inoculations per person per year and low (25%) or high (60%) levels of access to treatment.

**Supplementary Figure 25:**
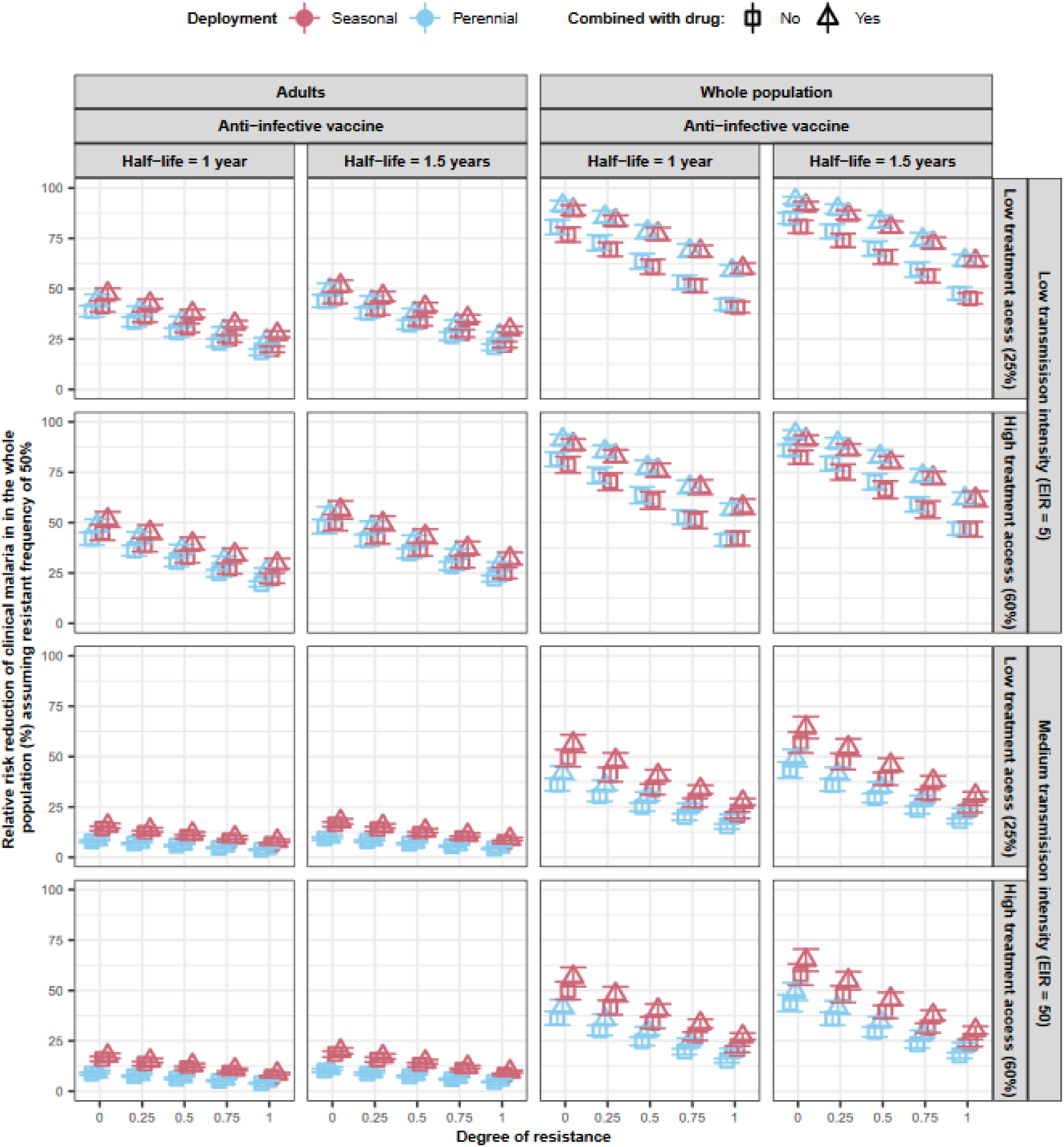
The impact of the degree of resistance on the effectiveness of vaccines that reduce transmission. Estimated vaccine effectiveness (VE) of the vaccine in a population composed of 50% sensitive genotypes and 50% resistant genotypes with different degrees of resistance (0, 0.25, 0.5, 0.75, 1) to vaccines **that aimed to reduce transmission**. The VE was estimated as the relative reduction in the number of clinical malaria cases across the entire population. The VE and 95% confidence intervals were predicted for each vaccine type and deployment strategy (perennial deployment (light red marker), seasonal deployment (light blue marker), with a drug (triangle), or without a drug (square) implemented with the vaccines. We evaluated the VE for vaccines with an initial efficacy of 95% and a half-life of 1.0 or 1.5 years. These vaccines were deployed with a coverage of 90% that did not decrease at the booster doses in settings with an EIR of 5 or 50 inoculations per person per year and low (25%) or high (60%) levels of access to treatment.

**Supplementary Figure 26:**
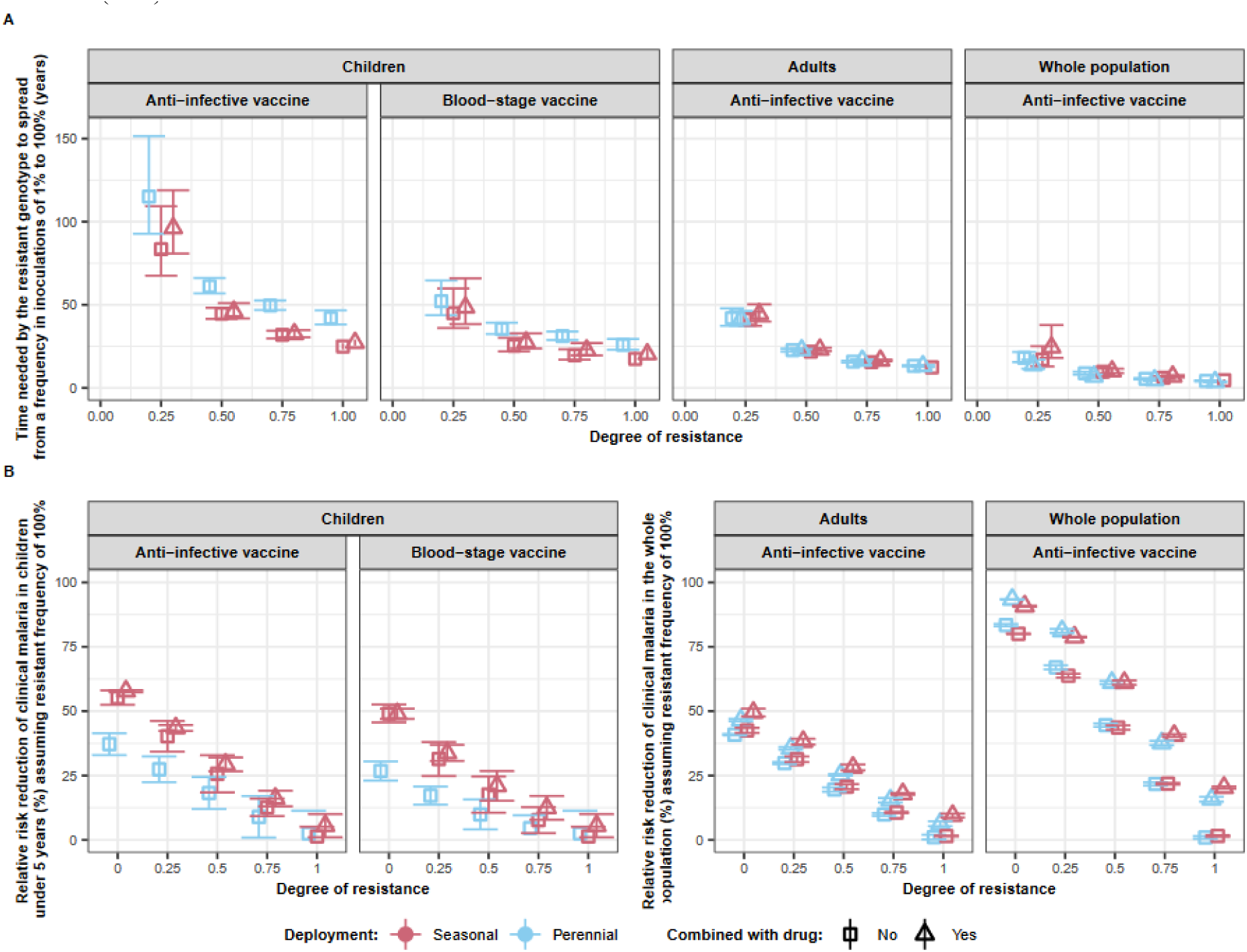
The impact of the degree of resistance on time needed before the resistant genotypes become fixed (100%) and vaccine effectiveness. (**A**) The predicted time needed for genotypes with different degrees of resistance (0, 0.25, 0.5, 0.75, 1) to vaccines to spread from a frequency in inoculations of 1% to 100%, *T_100_*. We predicted the *T_100_* and 95% confidence intervals for each vaccine type and deployment strategy (perennial deployment (light red marker), seasonal deployment with (light blue marker), with (triangle), or without (square) a drug implemented with the vaccines. (**B**) Estimated vaccine effectiveness (VE) and 95% confidence intervals of the vaccine in a population composed of 100% of resistant genotypes with different degrees of resistance (0, 0.25, 0.5, 0.75, 1) to the vaccine. For vaccines deployed to children, the VE was estimated as the relative reduction in the number of clinical malaria cases in children under five years. For the AIV deployed to the whole population or adults, the VE was estimated as the relative reduction in the number of clinical malaria cases in the whole population. The VE was predicted for various deployment strategies (perennial deployment (light red light marker), seasonal deployment with (light blue marker), with a drug (triangle), or without a drug (square) implemented with the vaccines. For (**A**) and (**B**), we assumed that the vaccine had an initial efficacy of 95%, a half-life of 1.5 years, and was deployed with a coverage of 90% that did not decrease at the booster doses in a setting with an EIR of 5 inoculations per person per year, and low levels of access to treatment (25%).

**Supplementary Figure 27:**
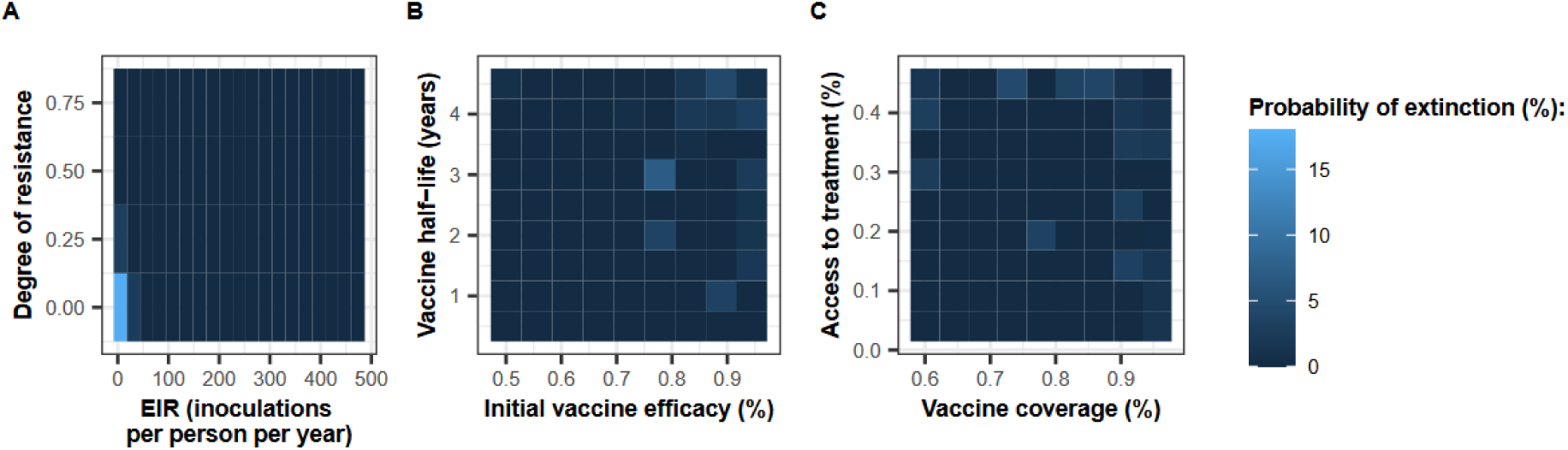
Probability of extinction across the explored parameter space. The figure illustrates the probability of extinction (defined as a prevalence of *P. falciparum* infection in the population of less than 0.1%) across the parameter space. This probability was estimated based on simulations that were run during the global sensitivity analysis. In these simulations, an AIV was deployed without a drug at constant coverage to the whole population in the seasonal setting. The probability of extinction (%) was estimated across the parameter range of (**A**) the EIR and degree of resistance, (**B**) initial vaccine efficacy and half-life, and (**C**) vaccine coverage and access to treatment.

#### 2.3 Estimation of the effectiveness of the vaccine

For each vaccine type and deployment strategy, we assessed the impact of the spread of a genotype resistant to the vaccine on the vaccine effectiveness (VE) against uncomplicated malaria episodes. To accomplish this, we calculated the VE in parasite populations where 50% were the sensitive genotype and 50% were the resistant genotype, with different degrees of resistance (0, 0.25, 0.5, 0.75) to the vaccine.

For vaccines that reduce morbidity, we defined the VE as the relative reduction of incidence of clinical malaria cases in children under five years during the year following the time at which the first cohort of vaccinated children received their last vaccine booster doses compared to the incidence of clinical malaria the year before the vaccine implementation (Supplementary Fig. 5). For vaccines that reduce the transmission, we assessed the VE as the relative reduction of incidence of clinical malaria cases in the whole population during the two years following vaccine implementation compared to the incidence of clinical malaria cases during the two years before vaccine implementation (Supplementary Fig. 6). The VE was estimated as follow,

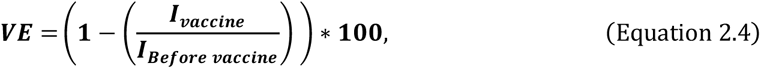

where *I_vaccine_* is the incidence rate (per person per month) of uncomplicated malaria episodes in the interested age group during the interested time, and *I_before_* vaccine is the incidence rate of uncomplicated malaria in the same age group during the same period of time but in the year(s) before vaccine deployment (Supplementary Fig. 5 and 6). The incidence rates (I) were calculated as,

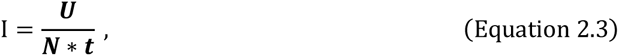

Where *U* is the number of uncomplicated malaria episodes that occur during the time of interest, *t* (*t*=1 years for vaccines preventing morbidity, and *t*=2 years for vaccines preventing transmission).

For each vaccine type and deployment strategy, we evaluated the VE when the resistant genotype had different degrees of resistance to vaccines. Vaccines had different half-lives (1 or 1.5 years) and an initial efficacy of 95%. We estimated the VE for the vaccine deployment at 90% coverage in a transmission setting with different transmission intensities (5 or 50 inoculations per person) and levels of access to treatment (25% or 60%). Each parameter combination was simulated in three stochastic realisations using a human population of 10,000 individuals and starting with a burn-in phase of 130 years before vaccine deployment. All the results can be seen in Fig. 3B and Supplementary Fig. 24 and 25. Supplementary Fig. 26B also display the results when we calculated the VE in parasite populations where 100% were the resistant genotype.

### 3 Supplementary Note 3: Supplementary results figure

